# Testing Denmark: A Danish nationwide surveillance study of COVID-19

**DOI:** 10.1101/2021.08.10.21261777

**Authors:** Kamille Fogh, Jarl E Strange, Bibi FSS Scharff, Alexandra RR Eriksen, Rasmus B Hasselbalch, Henning Bundgaard, Susanne D Nielsen, Charlotte S Jørgensen, Christian Erikstrup, Jakob Norsk, Pernille Brok Nielsen, Jonas H Kristensen, Lars Østergaard, Svend Ellermann-Eriksen, Berit Andersen, Henrik Nielsen, Isik S Johansen, Lothar Wiese, Lone Simonsen, Thea K. Fischer, Fredrik Folke, Freddy Lippert, Sisse R Ostrowski, Thomas Benfield, Kåre Mølbak, Steen Ethelberg, Anders Koch, Ute Wolff Sönksen, Anne-Marie Vangsted, Tyra Grove Krause, Anders Fomsgaard, Henrik Ullum, Robert Skov, Kasper Iversen

## Abstract

**Background:** National data on the spread of SARS-CoV-2 infection and knowledge on associated risk factors are important for understanding the course of the pandemic. “Testing Denmark” is a national large-scale epidemiological surveillance study of SARS-CoV-2 in the Danish population.

**Methods:** Between September and October 2020, approximately 1.3 million of 5.8 million Danish citizens (age > 15 years) were randomly invited to fill in an electronic questionnaire covering COVID-19 exposures and symptoms. The prevalence of SARS-CoV-2 antibodies was determined by Point-of Care rapid Test (POCT) distributed to participants home addresses.

**Findings:** In total 318,552 participants (24.5% invitees) completed the questionnaire and provided the result of the POCT. Of these, 2,519 (0.79%) were seropositive (median age 55 years) and women were more often seropositive than men, interquartile range (IQR) 42-64, 40.2% males. Of participants with a prior positive Polymerase Chain Reaction (PCR) test (n=1,828), 29.1% were seropositive in the POCT. Seropositivity increased with age irrespective of sex. Elderly participants (>61 years) reported less symptoms and had less frequently been tested for SARS-CoV-2 compared to younger participants. Seropositivity was associated with physical contact with SARS-CoV-2 infected individuals (Risk ratio (RR) 7.43, 95% CI: 6.57-8.41) and in particular household members (RR 17.70, 95% CI: 15.60-20.10). Home care workers had a higher risk of seropositivity (RR 2.09 (95% CI: 1.58-2.78) as compared to office workers. Geographic population density was not associated to seropositivity. A high degree of compliance with national preventive recommendations was reported (e.g., > 80% use of face masks), but no difference was found between seropositive and seronegative participants.

**Interpretation:** This study provides insight into the immunity of the Danish population seven to eight months after the first COVID-19 case in Denmark. The seroprevalence was lower than expected probably due to a low sensitivity of the POCT used or due to challenges relating to the reading of test results. Occupation or exposure in local communities were major routes of infection. As elderly participants were more often seropositive despite fewer symptoms and less PCR tests performed, more emphasis should be placed on testing this age group.

## Introduction

National seroprevalence data on antibodies to SARS-CoV-2 can guide national health policies in understanding transmission routes and thereby improve the management of potential new outbreaks during the COVID-19 pandemic (1–4). However, a large sample size is required to describe the spread of infection, risk factors, and severity of the infection across geography and demography (5).

Denmark has 5.8 million inhabitants (6) and as of July 5, 2021, there have been more than 295,654 (5 %) confirmed cases of SARS-CoV-2 infection and more than 2,537 COVID-19 related deaths in Denmark (7). The first confirmed case of SARS-CoV-2 infection in Denmark was reported on February 27, 2020 (8). In Denmark, the epidemic has been characterized by two infection waves; spring 2020 and autumn-winter 2020/2021, similar to several other European countries (9). Two lockdowns were imposed by the government, the first between March 11th to April 15th, 2020 and the second December 17th, 2020 to February 8th, 2021 (10). Testing for SARS-CoV-2 in Denmark using Polymerase Chain Reaction (PCR) was established in March 2020 and has been upscaled during the pandemic. From March 12 to April 21, 2020 individuals with moderate to severe symptoms of respiratory tract infection were offered testing. From April 21, 2020 testing was available for individuals with mild symptoms and asymptomatic contacts, and since May 18, 2020, nationwide high-intensity, free of charge testing for SARS-CoV-2 infection has been performed using PCR (11). Vaccination against COVID-19 began on December 27^th^, 2020 with residents and employees at nursing homes and frontline staff at hospitals (12).

The rates of COVID-19-related morbidity and mortality have been low in Denmark compared to other European countries (13). Nevertheless, considering the unknown proportion of asymptomatic or mildly symptomatic SARS-CoV-2 infected persons who have not been PCR-tested (viral throat- and nasopharyngeal swab), the population exposure to infection might be higher than reported according to PCR test findings (1). The seroprevalence has been reported for different groups in Denmark; blood donors (14), medical students (15), health care workers (16), a smaller national random selection of the population (10), homeless persons and sex workers (17) and persons from social housing areas (18), but hitherto no national investigation of this scale has been performed in Denmark.

“Testing Denmark” was a nationwide surveillance study of SARS-CoV-2 infection in the Danish population, launched in September 2020. The study was divided in 2 phases; phase 1 (the general population) and phase 2 (subgroups). In this article, we describe the process and results of phase 1. Results from phase 2 have been described elsewhere (17, 18).

The aim of this study was to explore possible risk factors for seropositivity by questionnaire data, and to determine the distribution of SARS-CoV-2 antibodies among Danish citizens over the age of 15 years, by the use of Point-of care rapid test (POCT) for antibodies against SARS-CoV-2.

## Methods

### Study design and participation

1.3 million Danish citizens over the age of 15 years (22 % of the population) were randomly drawn from the Civil Registration System (19) and invited to participate via the governmental, personal, password- protected digital mailbox system (e-Boks) from September 25, 2020. Written information about the project was available in 3 different languages; Danish, English and Arabic.

Participants were invited to complete a web-based questionnaire by a link (Enalyzer, Copenhagen, Denmark) in the invitation letter. The questionnaire included demographics, history of symptoms compatible with COVID-19, co-morbidities and substance use (see Appendix). In the questionnaire participants could further indicate if they wanted a POCT sent to their home address.

During October 2020 the POCT testing for SARS-CoV-2 IgG and IgM antibodies was performed by participants. Answers to the questionnaire and the result of the POCT results were registered by the participant in a secondary separate questionnaire sent to their e-Boks and returned to Enalyzer.

Detailed information about the test-procedure was provided with the invitation and could also be found at the project website (www.vitesterdanmark.dk), including instructional video on how to perform the POCT in practice, as well as videos with experts explaining different aspects of the study. Social media (Facebook and Instagram) were used for visualization. A call-center was set up for the participants to call in case of questions about the project, the questionnaire or how to perform the POCT.

Information about previous positive PCR test results amongst study participants was obtained from MiBa (The Danish Microbiological Database) that has complete coverage of all microbiological samples from general practice, hospitals and test facilities, analyzed by public laboratories (20).

### Detection of SARS-CoV-2 antibodies

The Livzon POCT (Livzon Diagnostics, Zhuhai, Guangdong, China) was used. The POCT is a lateral flow chromatographic immunoassay rapid test for qualitative detection and differentiation of anti-SARS-CoV-2 IgG and IgM antibodies in whole blood, which yields results in 15 minutes. The manufacturer reported a combined test sensitivity (either IgG or IgM positive) of 90.6% (95% CI: 86.6% - 93.4%) and a combined specificity (neither IgG nor IgM is positive) of 99.2% (95% CI: 97.6% - 99.7%) (21). An in-house validation (cases=150 individuals, controls=600 individuals) showed sensitivity of 93.3% and 92.7% and specificity of 98.2% and 97.5% for each of the two batches respectively (see appendix Table 4). The case panel samples were obtained from convalescent individuals within 2 months of disease onset. Sensitivity and specificity by self-use has not previously been studied.

The POCT was sent out with a small container of isotonic saline, capillary tubes, and fingerprickers. Participants were instructed by the use of the capillary tubes to add blood by fingerprick and isotonic saline to each of the two test cassettes (IgG and IgM). The test results were read by participants individually; positive results counted when both control line and test line appeared, the test was inconclusive when no control line appeared or if the reading chamber was discolored by blood. Inconclusive test results were treated as negative, as the test could not be repeated, participants only receiving one POCT for both IgG and IgM. Participants were categorized as seropositive if they had developed either IgG or IgM antibodies, or both against SARS-CoV-2.

### Outcome measures

The primary outcome of interest was to explore the association between SARS-CoV-2 infection, defined as a positive SARS-CoV-2 antibody self-test result (IgG and/or IgM), and putative risk factors for seropositivity.

The proportion of the study population with a positive antibody test for SARS-CoV-2 (as a proxy for previous infection) was a secondary outcome of interest.

### Approvals, ethics and registrations

This study was performed as a national surveillance study under the authority task of the national infectious disease control institute Statens Serum Institut (SSI), Copenhagen, Denmark. According to Danish law national surveillance activities from SSI do not require any individual approval from an ethics committee. The study was performed in agreement with the Helsinki II declaration and registered with the Danish Data Protection Authorities (P-2020-901). Participation was voluntary and all data were self- report ed. All personal data obtained in Enalyzer was kept in accordance with the general data protection regulation and data protection law stated by the Danish Data Protection Agency. Invitees received information about their legal rights and the use of their data in the invitation letter.

### Statistical analyses

Participants were considered seropositive if they tested positive for IgG, IgM, or both antibodies. Baseline characteristics of seropositive compared to seronegative persons are presented as numbers and percentages for categorical values and continuous values are presented as medians and interquartile ranges. The Wilcoxon rank test and chi-square test were used for comparisons of groups for continuous and categorial values, respectively. Unadjusted risk ratios (RR) with 95% confidence intervals (CI) were calculated for risk factors of seropositivity. We used logistic regression to calculate Odds ratios (OR) for seropositivity with 95% CI adjusted for sex, age, and household size for participants exposed to COVID-19 infected patients within the household. Data on population and areal by municipality for 2020 was obtained from Statistics Denmark (22). For participants with previous positive PCR test, we calculated the proportion of seropositive participants. Further, we analyzed the seroprevalence according to self-assed risk of being infected with SARS-CoV-2. Demographics were compared for responders and non-responders to the questionnaire. Further, demographics were compared for participants who provided the POCT results versus participants who did not provide the POCT results. P ˂0.05 was considered statistically significant. Data management, statistical analyses, and figures were performed and created using R version 3.2.1 (23).

## Results

### Baseline variables and association with seropositivity

In total, 474,411 participants (36.5% of invitees) replied to the electronic questionnaire and 397,843 received a POCT between October 2 and October 11, 2020. Invited persons who did not answer the questionnaire were more often males with lower participation among persons aged < 35 and > 74 years of age with no noticeable geographical variations (Supplementary table 1). Participants not providing POCT results were more often younger with no noticeable geographical variations (Supplementary table 1). The final study population comprised 318,552 participants (24.5% invitees) who answered the questionnaire and provided the results of the POCT (Figure 1). Age and sex distribution of the study population are shown in Supplementary figure 1.

**Figure 1:**
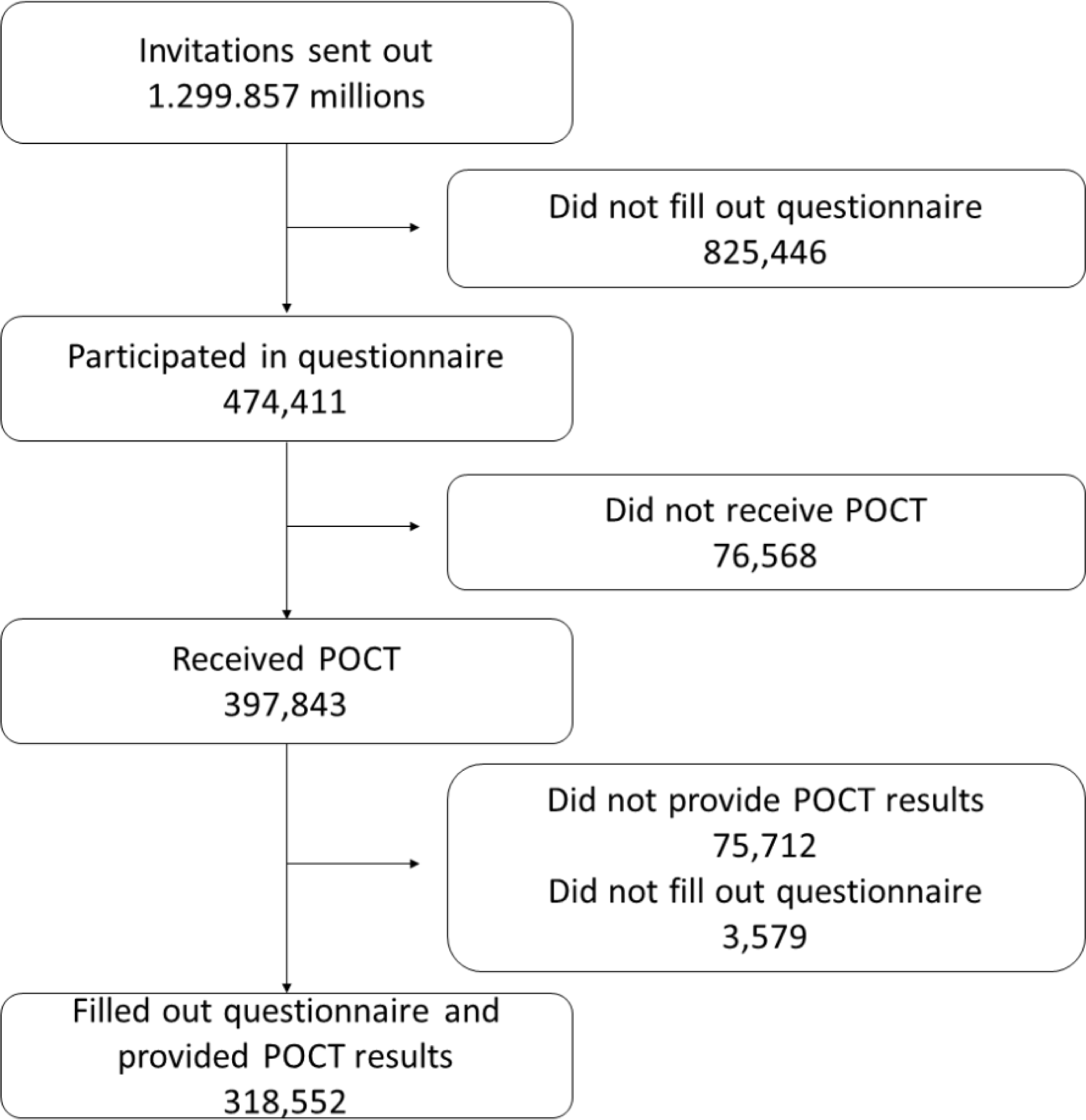
CONSORT diagram.

A total of 2,519 / 318,552 (0.79%) participants tested seropositive with 852 (0.27%) participants being positive for IgG antibodies, 1,078 (0.34%) for IgM antibodies, and 589 (0.18%) positive for both IgG and IgM antibodies. The seroprevalence increased with age with a higher proportion of IgM positive compared to IgG positive (age group 15-30: 0.24% IgG positive, 0.26% IgM positive, age group >61-75: 0.25% IgG positive, 0.38% IgM positive, Supplementary figure 2). No clear difference was found between IgG and IgM according to age groups (data not shown). For IgG, 9,294 (2.92%) and for IgM 9,269 (2.91%) were inconclusive, respectively.

Women were more likely to be seropositive (Table 1 and Supplementary figure 3). The comorbidity burden was higher in seropositive participants and reached statistical significance for participants with hypertension, stroke, diabetes, and chronic obstructive pulmonary disease (COPD). There was a numerically higher proportion of seropositive females among participants smoking >10 cigarettes per day and among participants consuming >21 standard drinks of alcohol per week. For body-mass index (BMI), the proportion of seropositive females was higher in the category underweight or obese, see Supplementary figure 4.

**Table 1:** Baseline characteristics of the study cohort on sex, age, BMI, smoking, alcohol use, previous test result and comorbidities stratified by seropositivity.

| Full cohort | Seronegative | Seropositive | p |
| --- | --- | --- | --- |
| **n** | 316,033 | 2,519 |  |
| **Age (years) (median [IQR])** | 53 [39-64]) | 55 [42-64] | 0.041 |
| **Male (%)** | 113,412 (42.2) | 1,012 (40.2) | <0.001 |
| **Body mass index (median [IQR])** | 25.4 [22.8, 28.7] | 25.5 [23, 29] | 0.115 |
| **Ever smoker (%)** | 168,024 (53.2) | 1,375 (54.6) | 0.161 |
| **Alcohol use\* (%)** | 36,747 (12.9) | 302 (13.5) | 0.443 |
| **Comorbidities (%)** |  |  |  |
| **Myocardial infarction** | 6562 (2.1) | 59 (2.3) | 0.389 |
| **Stroke** | 9067 (2.9) | 91 (3.6) | 0.030 |
| **Hypertension** | 82215 (26.0) | 711 (28.2) | 0.013 |
| **Diabetes** | 17528 (5.5) | 165 (6.6) | 0.032 |
| **Cancer** | 23250 (7.4) | 185 (7.3) | 1.000 |
| **Rheumatoid arthritis** | 19309 (6.1) | 176 (7.0) | 0.074 |
| **COPD** | 13872 (4.4) | 150 (6.0) | <0.001 |
| **Asthma** | 43996 (13.9) | 375 (14.9) | 0.172 |
| **Other chronic disease** | 56134 (17.8) | 456 (18.1) | 0.675 |

*\*Alcohol use: Reporting >7 units of alcohol a week for females or >14 units of alcohol for males*

### POCT findings in participants with previous COVID-19, or a positive PCR

When comparing self-estimated risk of infection with POCT results, only 0.5% of participants who self- estimated no prior infection were seropositive. Contrary, 13.5% of participants, who thought they had been infected, were seropositive. In comparison, 29.1% of participants, who had previously tested positive on a PCR test were seropositive (Supplementary figure 5 and Supplementary figure 6). For time between positive PCR test and POCT, 37.7 % were seropositive 21-30 days after the PCR test. The proportion of seropositive participants decreased with increasing time between PCR test and POCT (Supplementary figure 7). Differences between seropositive and seronegative who had previously tested positive on PCR test are shown in Table 3. Notably, time between positive PCR and POCT was lower for seropositive than seronegative participants.

**Table 3:** Characteristics of the study cohort who previously testes positive on PCR test.

| Full cohort | Seronegative | Seropositive | Total | p |
| --- | --- | --- | --- | --- |
| **n** | 1,296 | 532 | 1,828 |  |
| **Age (years) (median [IQR])** | 47 [31-59]) | 51 [40-61] | 49 [34-59] | <0.001 |
| **Male (%)** | 480 (37.0) | 233 (43.8) | 713 (39.0) | 0.008 |
| **Body mass index (median [IQR])** | 24.9 [22.4, 28.4] | 25.6 [23.0, 29.1] | 25.1 [22.6, 28.7] | 0.003 |
| **Days between pos. PCR and POCT (median [IQR])** | 58 [26, 188] | 38 [23, 176] | 46.5 [25, 187] | 0.082 |
| missing | 693 | 331 | 1,024 |  |
| **Comorbidities (%)** |  |  |  |  |
| Myocardial infarction | 26 (2.0) | 11 (2.1) | 37 (2.0) | 1.000 |
| Stroke | 31 (2.4) | 17 (3.2) | 48 (2.6) | 0.415 |
| Hypertension | 257 (19.8) | 129 (24.2) | 386 (21.1) | 0.041 |
| Diabetes | 67 (5.2) | 38 (7.1) | 105 (5.7) | 0.124 |
| Cancer | 75 (5.8) | 33 (6.2) | 108 (5.9) | 0.815 |
| Rheumatoid arthritis | 72 (5.6) | 31 (5.8) | 103 (5.6) | 0.907 |
| COPD | 46 (3.5) | 21 (3.9) | 67 (3.7) | 0.784 |
| Asthma | 202 (15.6) | 84 (15.8) | 286 (15.6) | 0.970 |
| Other chronic disease | 211 (16.3) | 84 (15.8) | 295 (16.1) | 0.850 |
| **Alcohol use\* (%)** | 144 (12.5) | 56 (11.7) | 22 (12.3) | 0.708 |
| **Ever smoker (%)** | 607 (46.8) | 278 (52.3) | 885 (48.4) | 0.040 |

Supplementary figure 8 and 9 shows geographical variations between municipalities in seropositivity and variations in population density. When ordering municipalities according to population density, no clear association between seropositivity and population density was found.

### Risk factors for seropositivity

#### Protective effect of authority recommendations

Most participants followed multiple recommended public health measures to prevent infection, e.g. >80% reported use of face masks. However, when examining serostatus according to behavior, no difference in serostatus was found between individual protective health measures, e.g. 82.5% of seronegative and 84% of seropositive reported use of face masks. (Figure 2).

**Figure 2:**
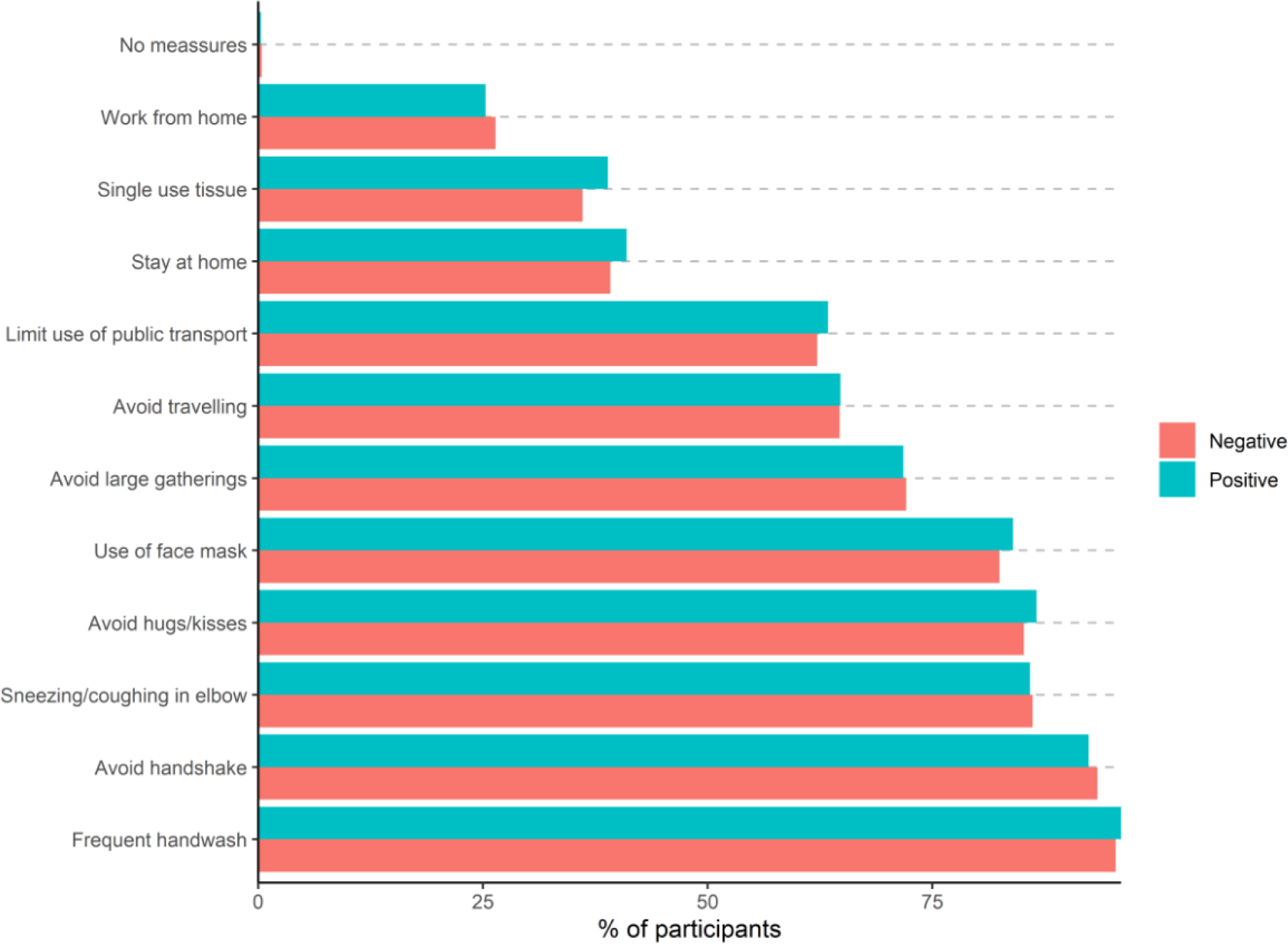
Proportion of participants following public health measures stratified for serostatus among 318,552 individuals.

#### Exposure to SARS-CoV-2 individuals

Participants who had physical contact or lived in a household with a SARS-CoV-2 infected person had the highest risk of being seropositive compared to participants who reported not to have been exposed to a SARS-CoV-2 infected person; RR of 7.43 (95% CI: 6.57 to 8.41) and 17.70 (95% CI: 15.60 to 20.10), respectively (Figure 3). Among participants exposed to a SARS-CoV-2 infected person within the household, the proportion of seropositive participants was higher in smaller household sizes (see Supplementary figure 10). However, when adjusting for sex, age, and household size, there was no significant increased risk for lower household size and risk of seropositivity (Table 2).

**Figure 3:**
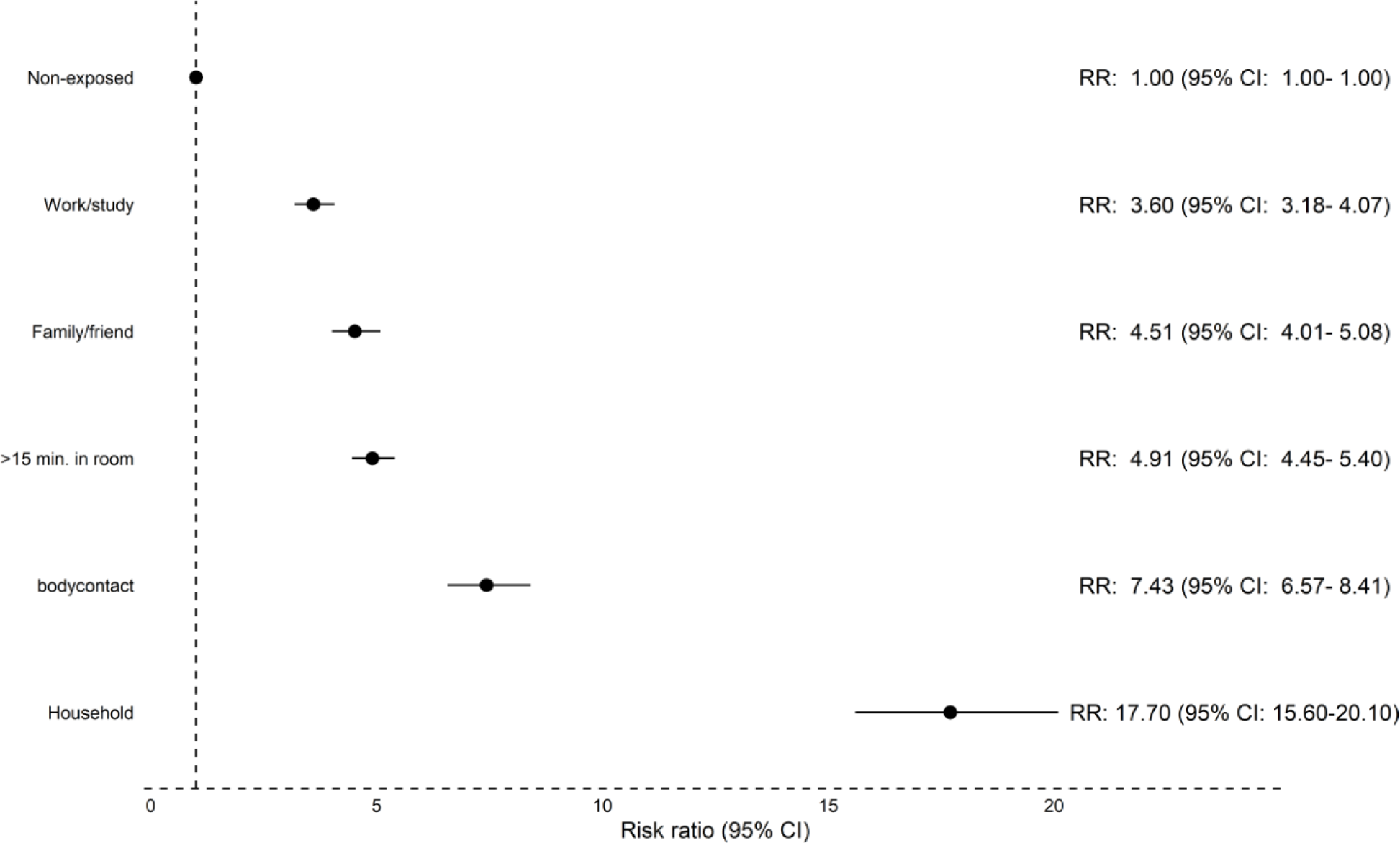
Risk ratio for seropositivity in a subset of 32,812 participants exposed to COVID-19 infected persons in various settings. For each setting, participants exposed to COVID-19 infected persons was compared to participants not exposed in this setting (reference group).

**Table 2:** Odds ratio for age, sex and household size stratified by seropositivity of the cohort

| Variable |  | Odds Ratio | 95% CI | p-value |
| --- | --- | --- | --- | --- |
| Age |  | 1.02 | [1.01;1.03] | <0.001 |
| Male |  | 1.01 | [0.77;1.34] | 0.920 |
| Household | 2 | Ref |  |  |
|  | 3 | 0.75 | [0.51;1.09] | 0.128 |
|  | 4 | 0.73 | [0.50;1.07] | 0.106 |
|  | 5 | 0.58 | [0.34;1.01] | 0.054 |
|  | >5 | 0.59 | [0.30;1.16] | 0.127 |

#### Occupation

Among professionals (full-time, part-time, and self-employed), working in the healthcare sector or with home care was associated with a higher risk of seropositivity compared to office work; healthcare sector: RR 2.02 (95%CI: 1.75 to 2.33), home care: RR 2.09 (95%CI: 1.58 to 2.78), see Figure 4.

**Figure 4:**
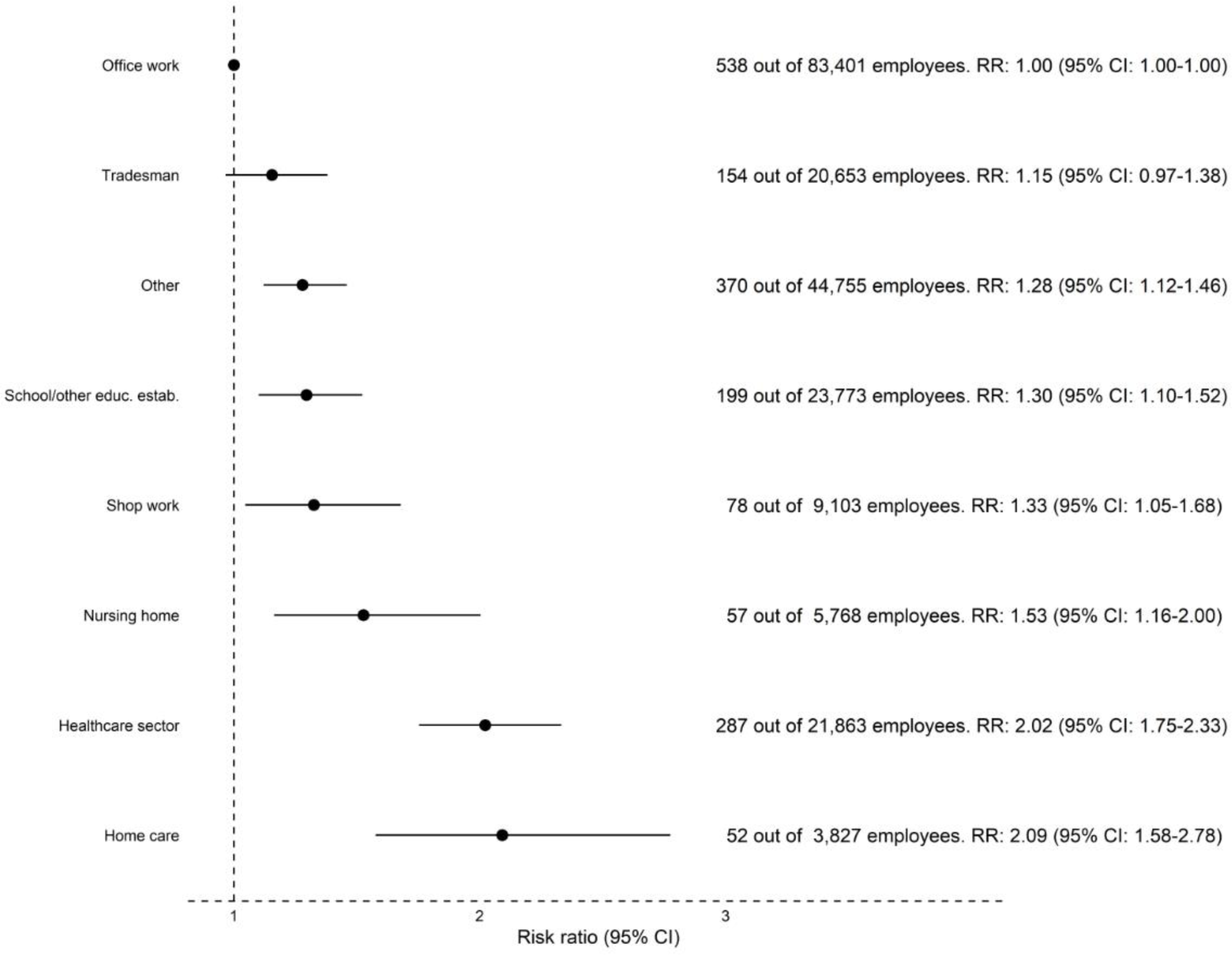
Risk ratio for seropositivity in a subset of 193,646 working (full-time, part-time, or self-employed) participants. Participants in each profession were compared to participants in office work.

#### Symptoms

For individual symptoms, loss of taste and smell were associated with the highest risk of being seropositive: ageusia (RR 5.91, 95% CI: 5.41 to 6.46) and anosmia (RR 4.84, 95% CI: 4.43 to 5.29). The risk of seropositivity for each symptom is shown in Figure 5.

**Figure 5:**
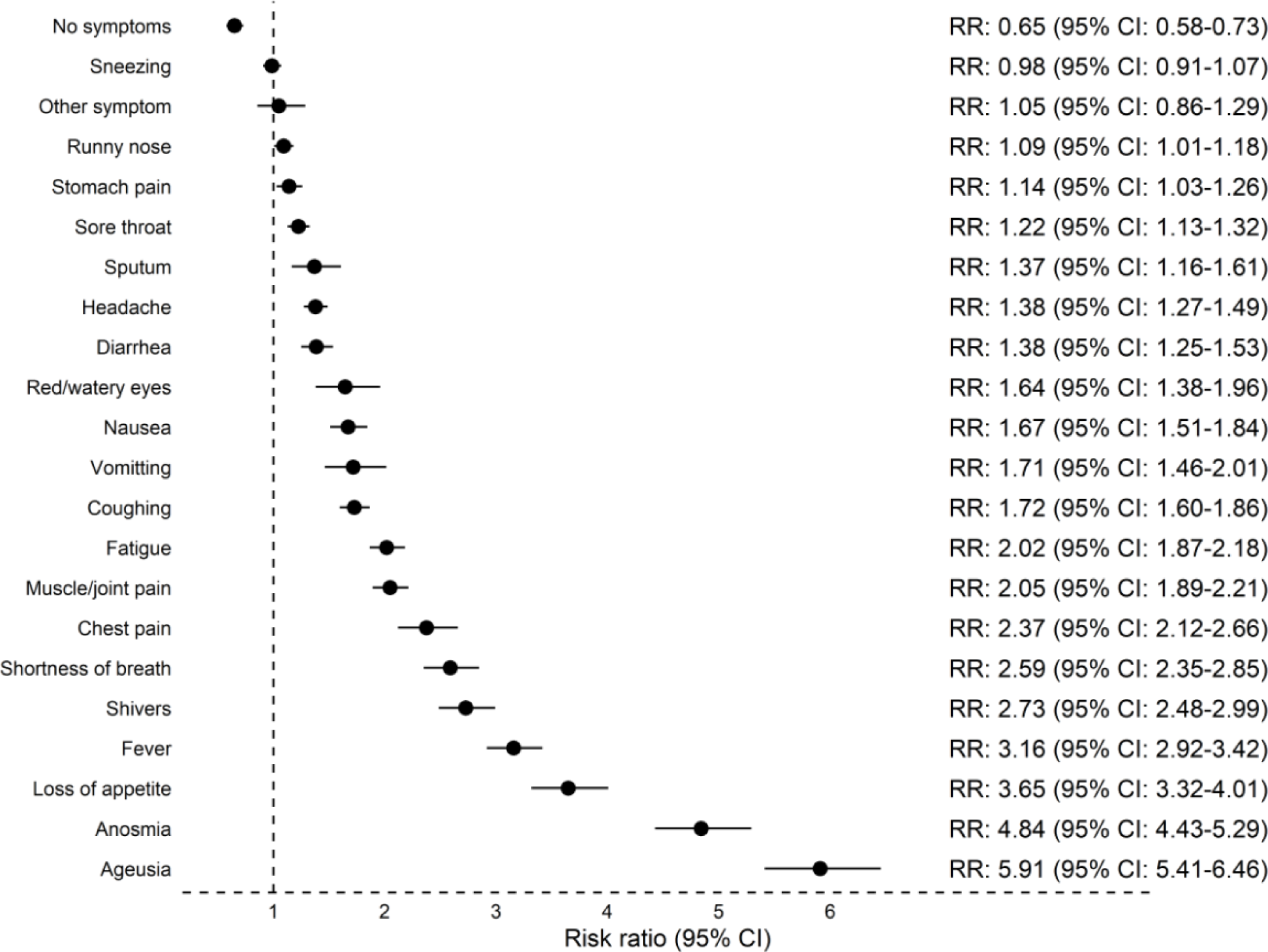
Risk of seropositivity for individual symptoms. Analysis included 318,552 participants.

Participants in advanced age groups had experienced less symptoms compared to participants in younger age groups with 39.5% in age group >75 years compared to 8.2% in the age group 15-30 years experiencing no symptoms (Supplementary figure 11). Further, participants in advanced age groups had been tested fewer times compared to participants in younger age groups irrespective of sex (Supplementary figure 12).

## Discussion

To our knowledge this is the largest population-based SARS-CoV-2 surveillance study performed. The main findings can be summarized as follows; females were found to have a higher seroprevalence than males.

Elderly participants were more often seropositive despite fewer symptoms and less often PCR tests. The geographical variation in seroprevalence was limited and did not seem to be related to population density. A prevalence of SARS-CoV-2 antibodies of only 0.79% was reported. Only 29% of PCR positive were POCT seropositive in our study. The study showed a high degree of adherence with national recommendation but there was no clear difference in reported compliance between seropositive and seronegative participants in the study period which covered the interval between the first infectious wave in spring 2020 and the second in autumn/winter.

### Age and sex

Until October 2020, 2.4 million people in Denmark had been tested with PCR at least once and up to multiple times, and 27,998 people were confirmed PCR positive (0.5% of the total population) (24). A population-based study in Denmark with 7,015 participants from August 2020 found a seroprevalence of 2.0% (age > 12 years) measured by Wantai SARS-CoV-2 Ab ELISA (10), the point estimates tended to be higher in the age group 18-39 years and lower in the age group >65 years, with no difference observed by sex. Also, a convenience sample of blood donors tested in October 2020 with ELISA found a seroprevalence of 2.1% (adults aged 18-70) (25). In contrast, we found a seroprevalence of only 0.79%, higher proportions of seropositivity in younger age groups and females being seropositive more often.

A Danish study of household transmission, with individual level register data on all national PCR test for SARS-CoV-2 for the period February-July 2020, suggested that susceptibility to infection increases with the age of the susceptible person (26). Other international studies tends to show trends in line with our results with an increase in seropositivity with age (9) and females having increased IgG positivity (27). By sending our test material to participants at home, we may have been able to include vulnerable and elderly susceptible to infection who otherwise would not have had the opportunity to participate. This is supported by our findings that participation in POCT was high in all age groups except the younger age group. This could partially explain the difference in seroprevalence between our study and aforementioned Danish studies, which included healthy blood donors as well as a population that should attend a venous blood sample.

### Testing and symptoms

In Denmark, one of the measures to contain the epidemic has been to offer easy-access, free of charge testing. The Danish Health authorities have encouraged the population to have test performed in case of symptoms of COVID-19 or after close contact with infected persons. Elderly participants reported fewer previous tests. When compared to younger participants, elderly participants might have fewer social contacts and/or could have isolated themselves to a higher degree thus avoiding potential close contact with infected persons. Further, younger participants may be more exposed to infection by having more social contacts or via their employment. However, in a recent report by the HOPE project (How Democracies Cope with COVID19), elderly people in Denmark were not found to report higher levels of self-quarantine when experiencing symptoms or when testing positive by PCR compared to younger people (28).

The strongest correlation to seropositivity was ageusia and anosmia (loss of taste and smell), consistent with previous findings (15, 16, 18). In general, we found that seropositive participants more frequently recalled having had symptoms when compared to seronegative participants.

When stratifying for age groups, elderly participants reported symptoms less frequently. It may be that only the healthiest elderly participated, however, this outcome could also be a bias resulting from comorbidity disorders and long recall period. Our results are surprising because aging itself has been associated with more severe COVID-19 symptoms due to increased comorbidities with age and more aggressive clinical behavior (29). Nevertheless, the level of antibodies (comparable levels of IgG and IgM) was highest among elderly participants although they reported fewer symptoms and had fewer tests. As such, elderly participants may more often be subject to asymptomatic infections, thereby constituting an important subgroup that may warrant further attention. However, it should also be noted that individuals in the working age who were unable to work from home may attend PCR testing more often than people who have retired, and this could contribute to our observations.

### Occupation

As previously reported, working in the health care sector was associated with a higher risk of seropositivity (16, 30). Working in home care or at nursing homes also increased the risk. These occupations often involves working with patients and being in close physical contact to other persons, thus increasing the risk of infection (30). The proportion of females working in the health care sector is typically higher than males (31), possibly explaining the higher proportion of seropositive females. Conversely, those who have office jobs, and therefor possibly better opportunities to work from home, have been at less risk of infection during the first infectious surge.

### Behavior and household

We observed a high proportion of participants following the authority’s recommendations to reduce the risk of SARS-CoV-2 infection. Remarkably, seropositive participants were slightly more compliant with these recommendations compared to seronegative participants on almost all the preventive measures. However, participants who are more attentive to recommendations, e.g. health care professionals are more exposed to SARS-CoV-2 infection. As such, the effect of the authority’s recommendations could be underestimated. Household composition is an important venues for transmission of infection due to household size and living conditions (32). Sustained close contact and crowded indoor environments pose a higher risk of transmission (33, 34). A metanalysis by Madewell et al indicated that household and family members are at higher risk of infection compared with other types of close contacts, and spouses were at higher risk compared with family contacts. Further, household crowding (e.g. number of people per room) may be more important for transmission than the total number of people per household (32). Our results showed seropositivity to be highest among smaller households with only two household members, possibly due to two person households often comprising of couples with close contact, and thereby increased risk of transmission. This finding is also consistent with a previous preprint study on SARS-CoV-2 transmission within Danish households, which demonstrated a transmission pattern that was exponentially decreasing with the number of members in the household (26).

### Strengths and limitations

This population-based study had a broad national participation with 22% of the population invited and a response rate of 36.5% among the invitees for the questionnaire and 24.5% for the POCT. To determine the distribution of infectious disease, serological surveys with a representative sample of the wider population are important, particularly in the presence of asymptomatic individuals or incomplete ascertainment of those with symptoms.

This study has limitations. Recruitment of participants by e-Boks might exclude the proportion of the population that are without or have only limited access to this digital governmental information system and less technology-proficient individuals, or marginalized groups who are seen to have a higher risk of infection (17, 18). A smaller proportion of residents may not have been able to read and understand Danish , English or Arabic. People under the age of 15 years were not included and the findings are not applicable to children. Data on ethnicity was not available from the questionnaire. The recall period of symptoms was long, up to 7 months. Information on the exact point of time for participants becoming infected or turning seropositive was not available. In addition, persons with a previous positive PCR may have been less inclined to participate, thereby resulting in selection bias and potentially underestimating the true seroprevalence. Conversely, particularly persons working in health care or nursing may have had an increased interest to know about possible protective immune status due to their working tasks and knowledge of former infection and/or increased exposure.

The low seroprevalence at 0.79% in our study may be due to low sensitivity of the POCT used or due to challenges relating to the reading of the test results, since 2.9% were inconclusive. POCT in general have a lower diagnostic performance compared to laboratory testing (35). Test results also depend on the prevalence of infection in the population which will be low when screening asymptomatic and higher for those with suggestive symptoms. In low prevalence settings, true positive test results are uncommon. As such, the predictive value of a positive test will be lower in individuals with a low background risk of infection (36). Only 0.5% of the Danish population were confirmed PCR positive during the study period. The diagnostic testing window is also of importance as the study was performed seven to eight months after the first COVID-19 case in Denmark. The antibody response of IgM and IgG is found to be highest about 2-3 weeks and 3-4 weeks, respectively after symptom onset and decrease afterwards (35). 37% of our study participants had a positive POCT 20-30 days after a positive PCR. In addition, we found that for seronegative, longer time had passed from a previously positive PCR test than for seropositive. As participants performed the POCT at home, incorrect testing procedure or misinterpreted POCT results could lead to false negative POCT results. Importantly, inconclusive tests were treated as negative in our study, and weak lines suggesting a positive test result, could be misinterpreted as a negative test result. In other Danish studies, the tests (POCT and ELISA) have been performed and read or analyzed by professional staff which increases the performance of the test. Consequently, the seroprevalence is likely underestimated in our study. However, seropositivity was low among participants who did not have a previous positive PCR test, indicating a high specificity of the POCT, thus the associations found are reliable.

### Perspectives

To date, this is the largest population-based Danish study where test material has been sent to participants and performed at home with broad national participation. Nationwide information can be difficult to gather and the study design in question presents a novel way for conducting future studies. Additionally, this setup can be used as a model for ongoing monitoring of COVID-19 immunity in the population, both from past infection and from vaccination against SARS-CoV-2.

## Conclusion

This study provides insight into the immunity of the Danish population seven to eight months after the first COVID-19 case in Denmark. The seroprevalence was lower than expected probably due to low sensitivity of the POCT used or due to challenges relating to the reading of test results. Future studies could be improved with an easier POCT test to perform and a shorter questionnaire. A high degree of compliance with national preventive recommendations was seen, but no clinically significant protective effect was identified.

Occupation, domestic exposure and other known exposures in the local communities were clear routes of infection, and in particular transmission in two person households, served as a major domain of transmission. As elderly participants were more often seropositive despite fewer symptoms and less testing, more emphasis should be placed on testing this age group.

## Contributors

The study was designed and initiated by: KF, BS, RS, HU and KI. Data analysis was done by: JS and KF

The first draft was written by: KF, JS, HB, RS and KI

All authors have critically revised the manuscript and agree to be accountable for all aspects of the work. All authors approved the final version of the manuscript.

## Data Availability

All personal data obtained in Enalyzer was kept in accordance with the general data protection regulation and data protection law stated by the Danish Data Protection Agency. The study was performed in agreement with the Helsinki II declaration and registered with the Danish Data Protection Authorities (P-2020-901).

## Acknowledgements

The authors would like to thank the Danish Ministry of Health, the Danish Patient Safety Authority, the Local Government Denmark, Danish Regions, Danish Patients, DaneAge Association, the Danish Medical Association, the Danish Nurses Organization, the Danish Heart Association, the Danish Cancer Society, the Danish Lung Association, the Danish National Organization for homeless people (SAND), the Danish Family Planning Association and the Council for Ethnic Minorities for support of the study.

## Supplementary materials

**Supplementary table 1:** Sex and age group distribution and place of living for responders and non- responders among population invited to participate in the questionnaire. Missing data represents invited persons who requested to be removed. Also shown are sex and age group distribution and place of living for participants who provided POCT results versus participants who did not provide the POCT results.

|  | Answered Questionnaire |  |  | Provided POCT result |  |  |
| --- | --- | --- | --- | --- | --- | --- |
| N<br>Row %<br>Col % | No | Yes | Total | No | Yes | Total |
| **Females** | 384,507<br>58.6 %<br>46.6 % | 272,012<br>41.4 %<br>57.3 % | 656,519<br>50.5 % | 43,538<br>18.9 %<br>54.4 % | 186,342<br>81.1 %<br>57.9 % | 229,880<br>57.2 % |
| **Males** | 440,939<br>68.5 %<br>53.4 % | 202,399<br>31.6 %<br>42.8 % | 643,338<br>49.5 % | 36,433<br>21.2 %<br>45.6 % | 135,789<br>78.9 %<br>42.2 % | 172,222<br>42.8 % |
| **Total** | 825,446<br>63.5 % | 474,411<br>36.5 % | 1,299,857<br>100.00 | 79,971<br>19.9 % | 322,131<br>80.1 % | 402,102<br>100 % |
| Age group, N<br>Row %<br>Col % | No | Yes | Total | No | Yes | Total |
| **15-25** | 147,686<br>77.4 %<br>17.9 % | 43,130<br>22.6 %<br>9.1 % | 190,816<br>14.7% | 10,122<br>28.7 %<br>12.7 % | 25,112<br>71.3 %<br>7.8 % | 35,234<br>8.8 % |
| **25-34** | 141,758<br>69.3 %<br>17.2 % | 62,679<br>30.7 %<br>13.2 % | 204,437<br>15.7 % | 15,014<br>27.9 %<br>18.8 % | 38,831<br>72.1 %<br>12.1 % | 53,845<br>13.4 % |
| **35-44** | 113,045<br>62.2 %<br>13.7 % | 68,857<br>37.9 %<br>14.5 % | 181,902<br>14.0 % | 14,382<br>24.1 %<br>18.0 % | 45,212<br>75.9 %<br>14.0 % | 59,594<br>14.8 % |
| **45-54** | 116,215<br>54.6 %<br>14.1 % | 96,645<br>45.4 %<br>20.4 % | 212,860<br>16.4 % | 16,488<br>19.8 %<br>20.6 % | 66,803<br>80.2 %<br>20.7 % | 83,291<br>20.7 % |
| **55-64** | 99,021<br>50.1 %<br>12.0 % | 98,632<br>49.9 %<br>20.8 % | 197,653<br>15.2 % | 13,581<br>16.1 %<br>17.0 % | 70,831<br>83.9 %<br>22.0 % | 84,412<br>21.0 % |
| **65-74** | 94,985<br>55.7 %<br>11.5 % | 75,551<br>44.3 %<br>15.9 % | 170,536<br>13.1 % | 7,332<br>11.6 %<br>9.2 % | 56,041<br>88.4 %<br>17.4 % | 63,373<br>15.8 % |
| **75+** | 112,736<br>79.6 %<br>13.7 % | 28,917<br>20.4 %<br>6.1 % | 141,653<br>10.9 % | 3,052<br>13.7 %<br>3.8 % | 19,301<br>86.4 %<br>6.0 % | 22,353<br>5.6 % |
| **Total** | 825,446<br>63.5 % | 474,411<br>37.0 % | 1,299,857<br>100 % | 79,971<br>19.89 | 322,131<br>80.11 | 402,102<br>100.0 % |
| Place of living, N<br>Row %<br>Col % | No | Yes | Total | No | Yes | Total |
| **The Capital**<br>**Region of**<br>**Denmark** | 262,620<br>63.9 %<br>31.8 % | 148,622<br>36.1 %<br>31.3 % | 411,242<br>31.6 % | 27,798<br>21.9 %<br>34.8 % | 99,008<br>78.1 %<br>30.7 % | 126,806<br>31.5 % |
| **Region Zealand** | 121,320<br>64.6 %<br>14.7 % | 66,595<br>35.4 %<br>14.0 % | 187,915<br>14.5 % | 11,779<br>21.0 %<br>14.7 % | 44,334<br>79.0 %<br>13.8 % | 56,113<br>14.0 % |
| **The Region of**<br>**Southern**<br>**Denmark** | 174,432<br>63.9 %<br>21.1 % | 98,359<br>36.1 %<br>20.7 % | 272,791<br>21.0 % | 15,338<br>18.5 %<br>19.2 % | 67,761<br>81.5 %<br>21.0 % | 83,099<br>20.7 % |
| **Central Denmark**<br>**Region** | 183,623<br>62.1 %<br>22.3 % | 111,950<br>37.9 %<br>23.6 % | 295,573<br>22.7 % | 17,313<br>18.3 %<br>21.7 % | 77,292<br>81.7 %<br>24.0 % | 94,605<br>23.5 % |
| **The North**<br>**Denmark Region** | 83,451<br>63.1 %<br>10.1 % | 48,885<br>36.9 %<br>10.3 % | 132,336<br>10.2 % | 7,743<br>18.7 %<br>9.7 % | 33,736<br>81.3 %<br>10.5 % | 41,479<br>10.3 % |
| **Total** | 825,446<br>63.5 % | 474,411<br>36.5 % | 1,299,857<br>100 % | 79,971<br>19.89 | 322,131<br>80.1 % | 402,102<br>100.0 % |
| Missing 21 |  |  |  |  |  |  |

**Supplementary figure 1:**
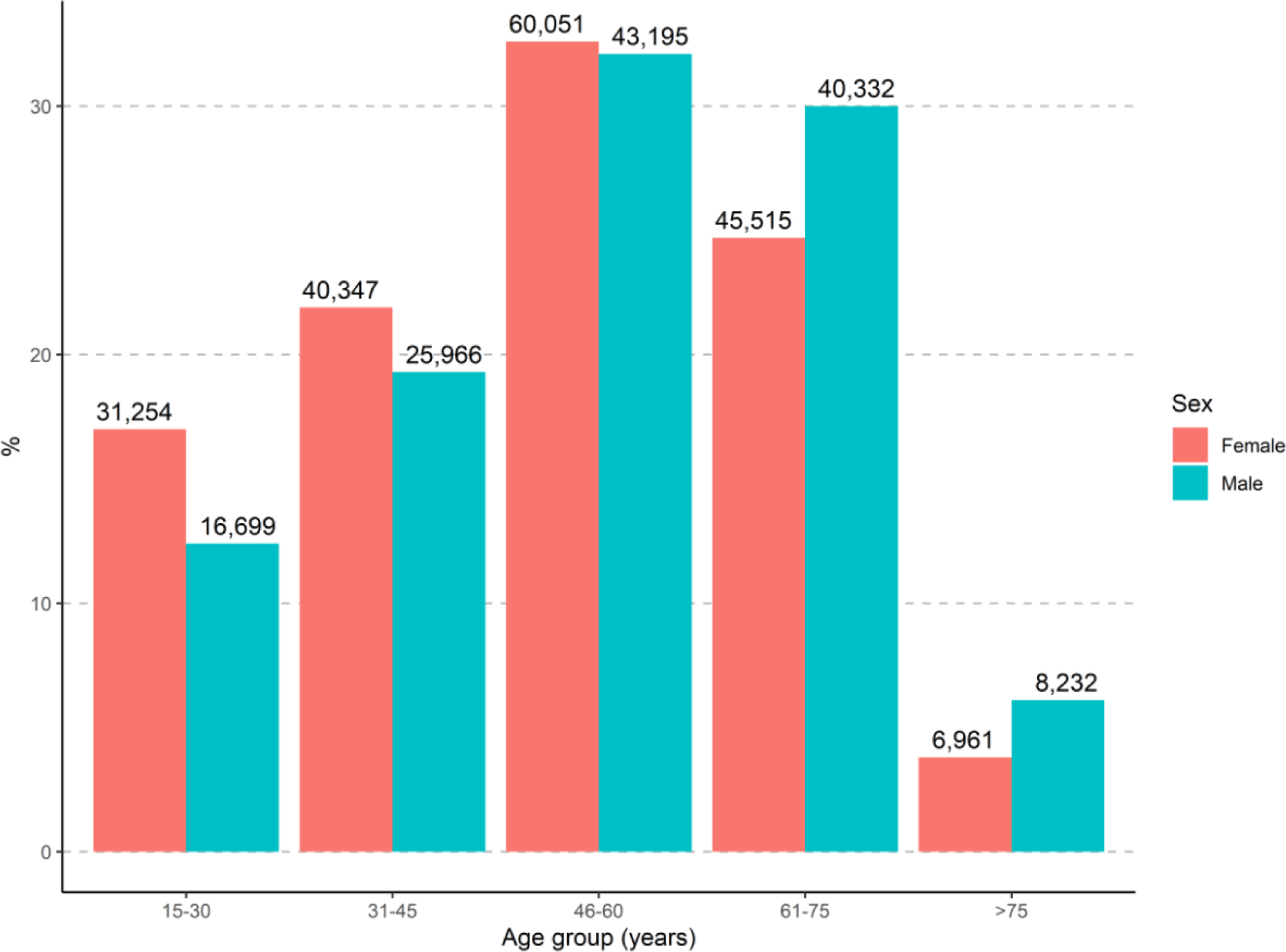
Age and sex distribution of the final study population. Numbers above bars represent number of participants in each group.

**Supplementary figure 2:**
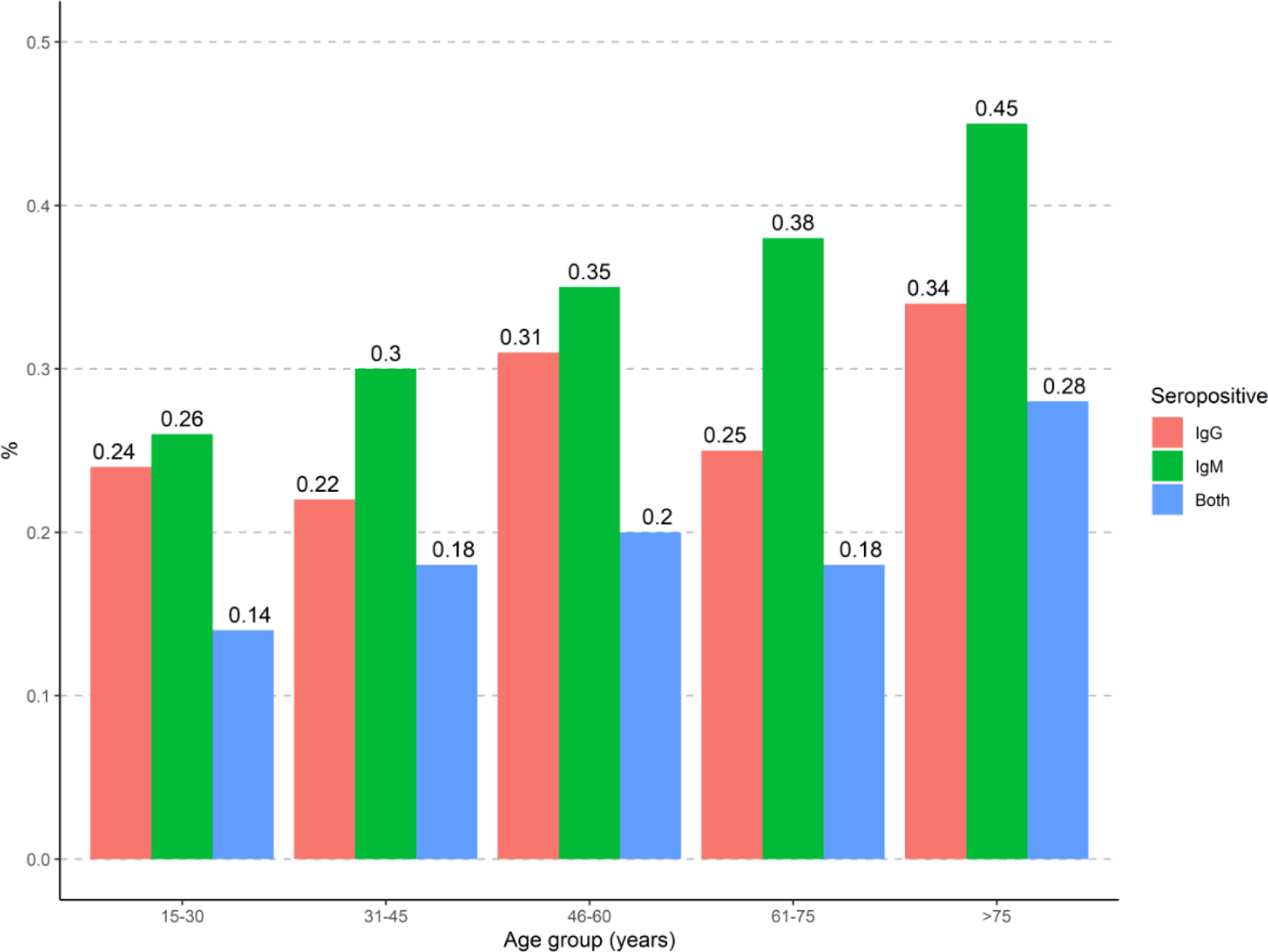
Distribution of SARS-CoV-2 antibodies according to age groups. Numbers above bars represent percentage of total number of participants in groups.

**Supplementary figure 3:**
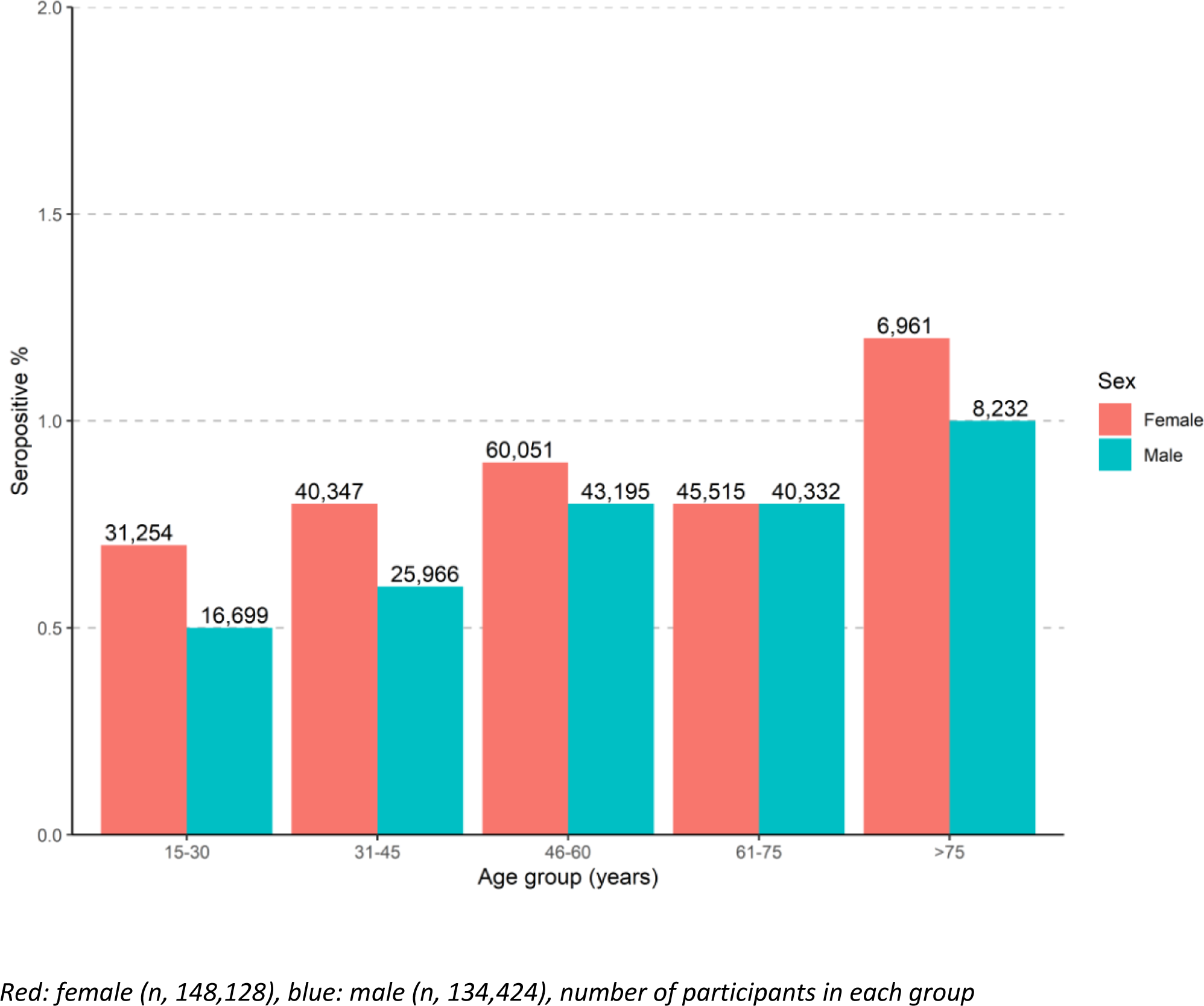
SARS-CoV-2 seropositive % among 318,552 individuals stratified for age groups and sex. Numbers above bars represent total of participants within each group.

**Supplementary figure 4:**
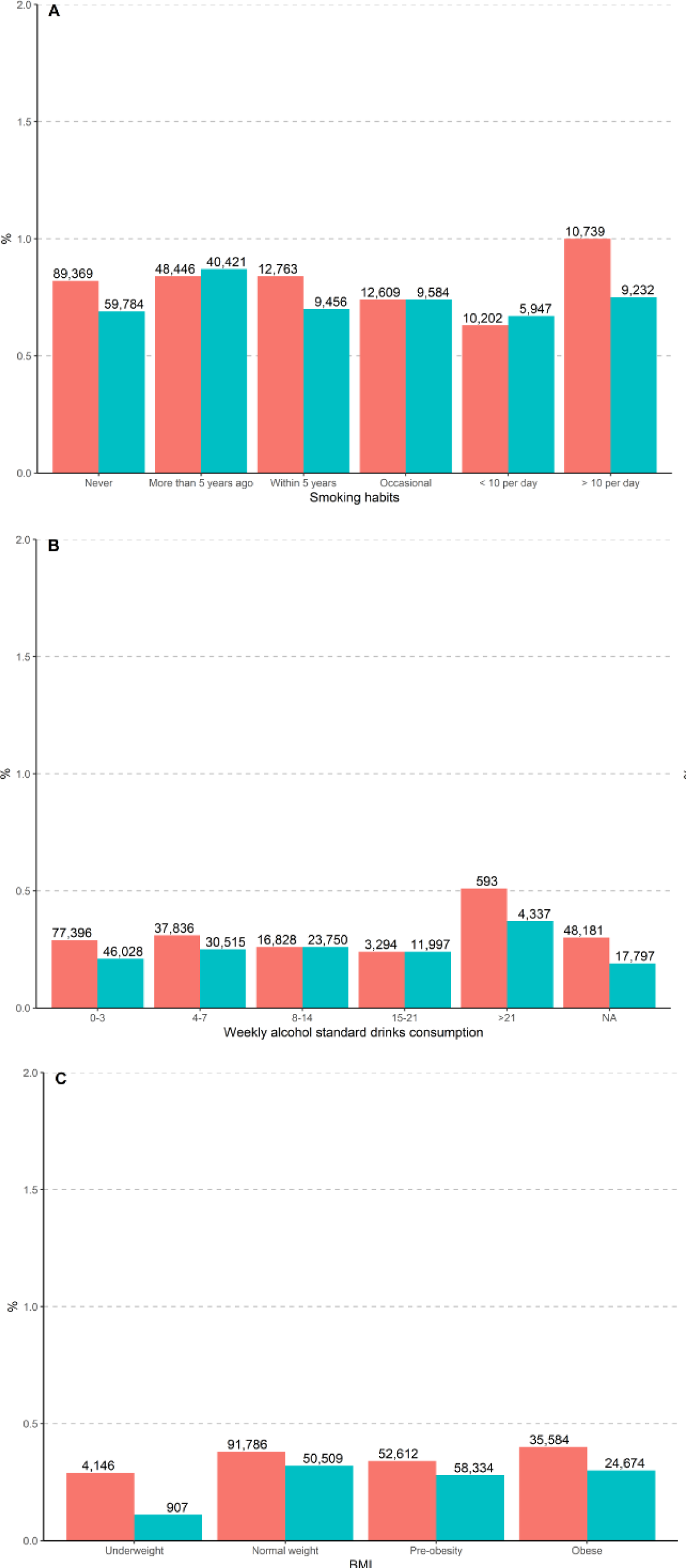
SARS-CoV-2 seropositive % according to smoking habits, weekly alcohol consumption, and BMI stratified for sex. Red bar represents females, blue bar represents males. Numbers above bars represent number of participants in each group.

**Supplementary figure 5:**
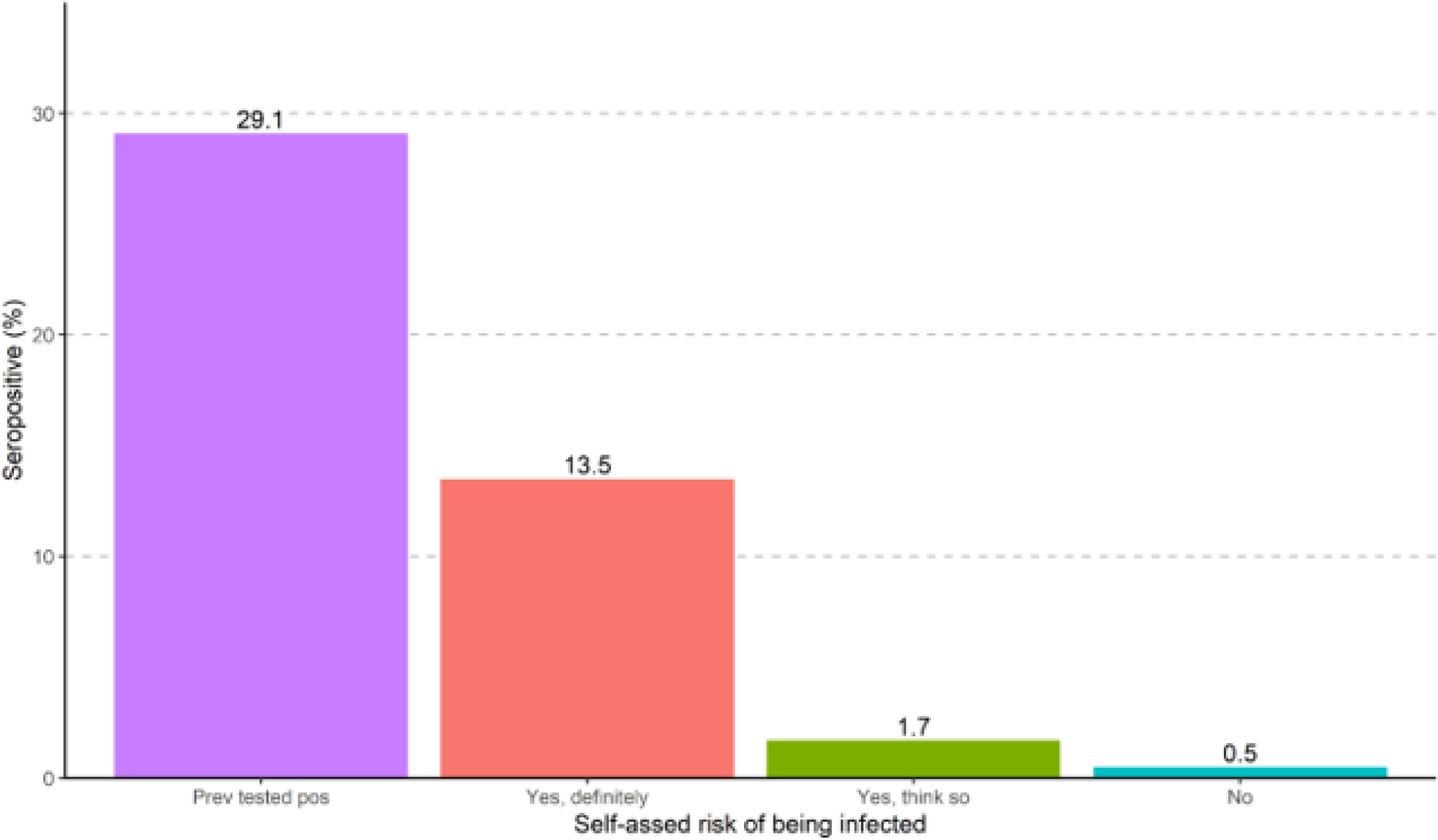
Seropositive % for self-assed risk of being infected and seropositive % for participants with a previous positive PCR test. Purple bar represents a subset of 1,828 participants with a positive PCR test prior to POCT test. Red, green, and blue bars represent participants without a prior positive PCR test. The self-assed risk of being infected was compared to the result of POCT test

**Supplementary figure 6:**
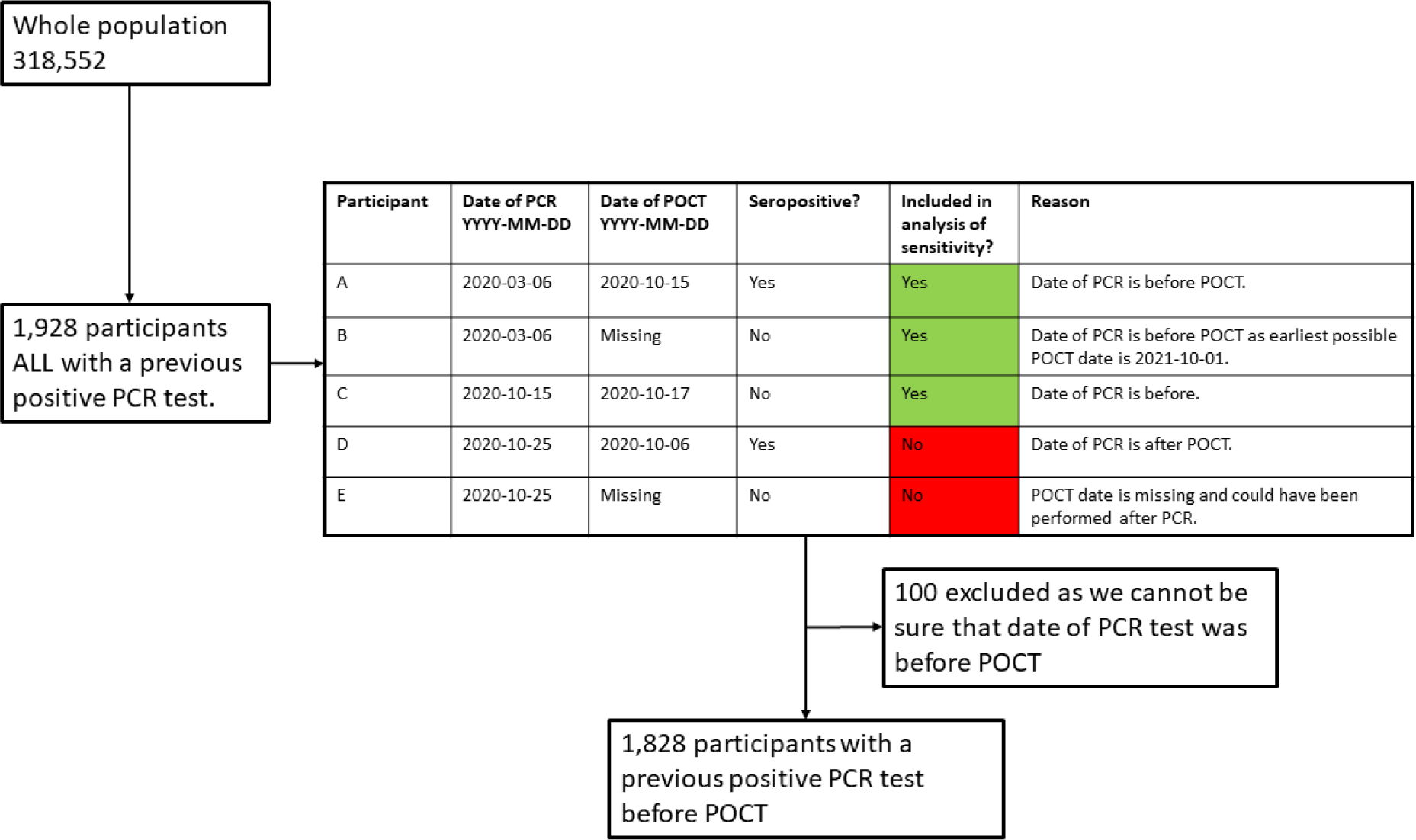
Flowchart for identifying participants with a positive PCR test before POCT.

**Supplementary figure 7:**
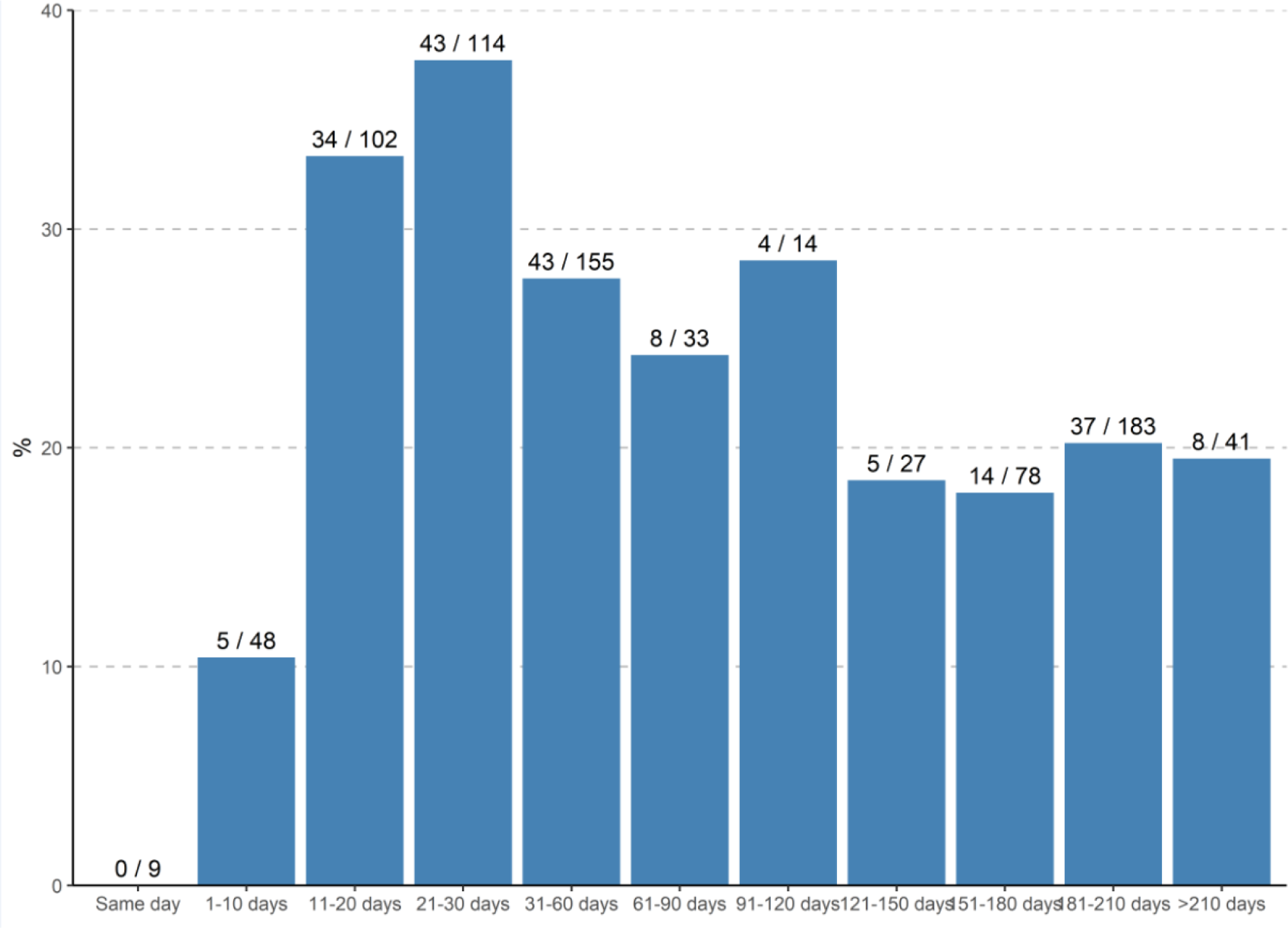
SARS-Cov-2 seropositive % among 804 individuals by POCT stratified for days since positive PCR.

**Supplementary figure 8:**
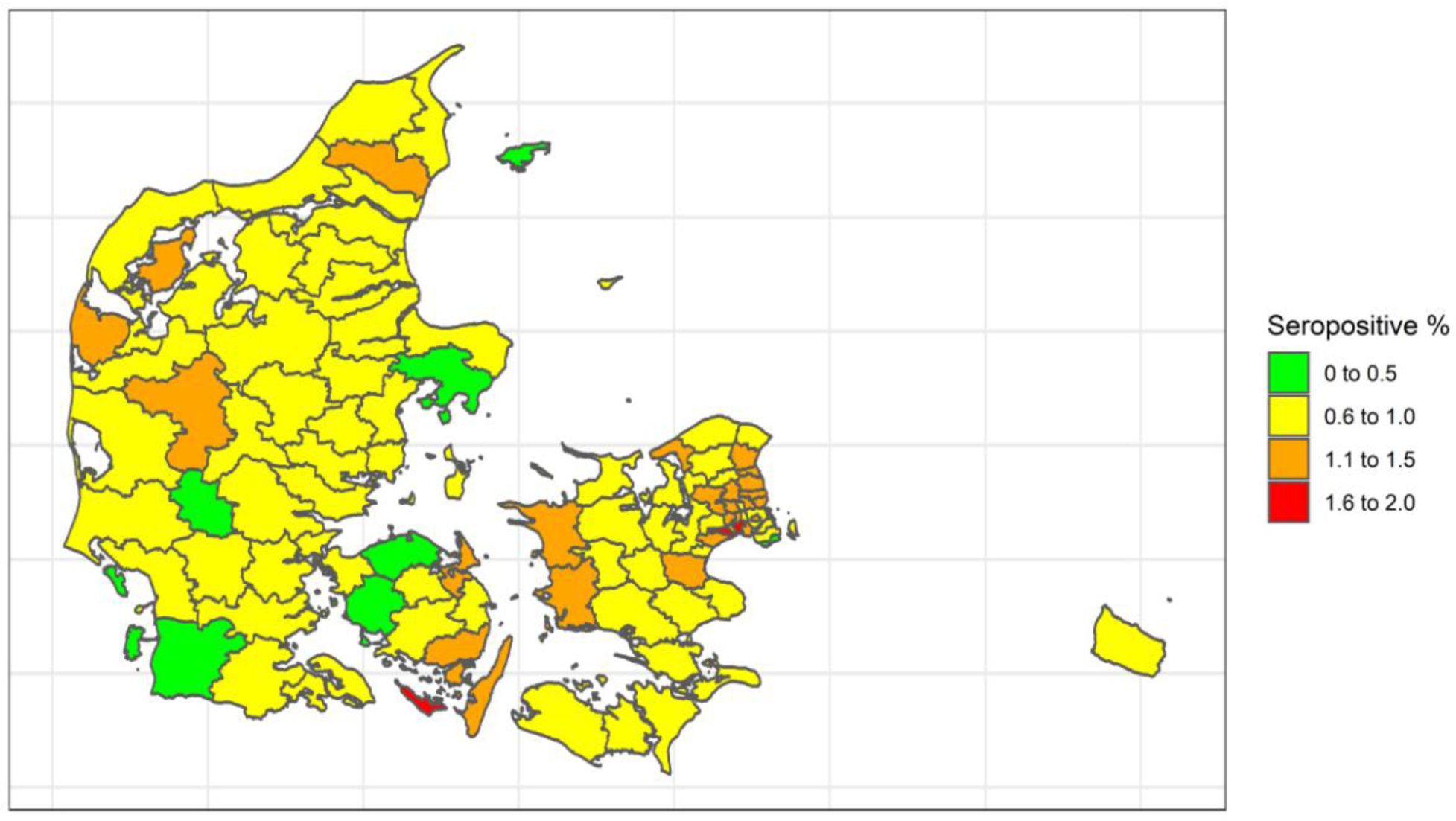
Map of seropositivity for each municipality.

**Supplementary figure 9:**
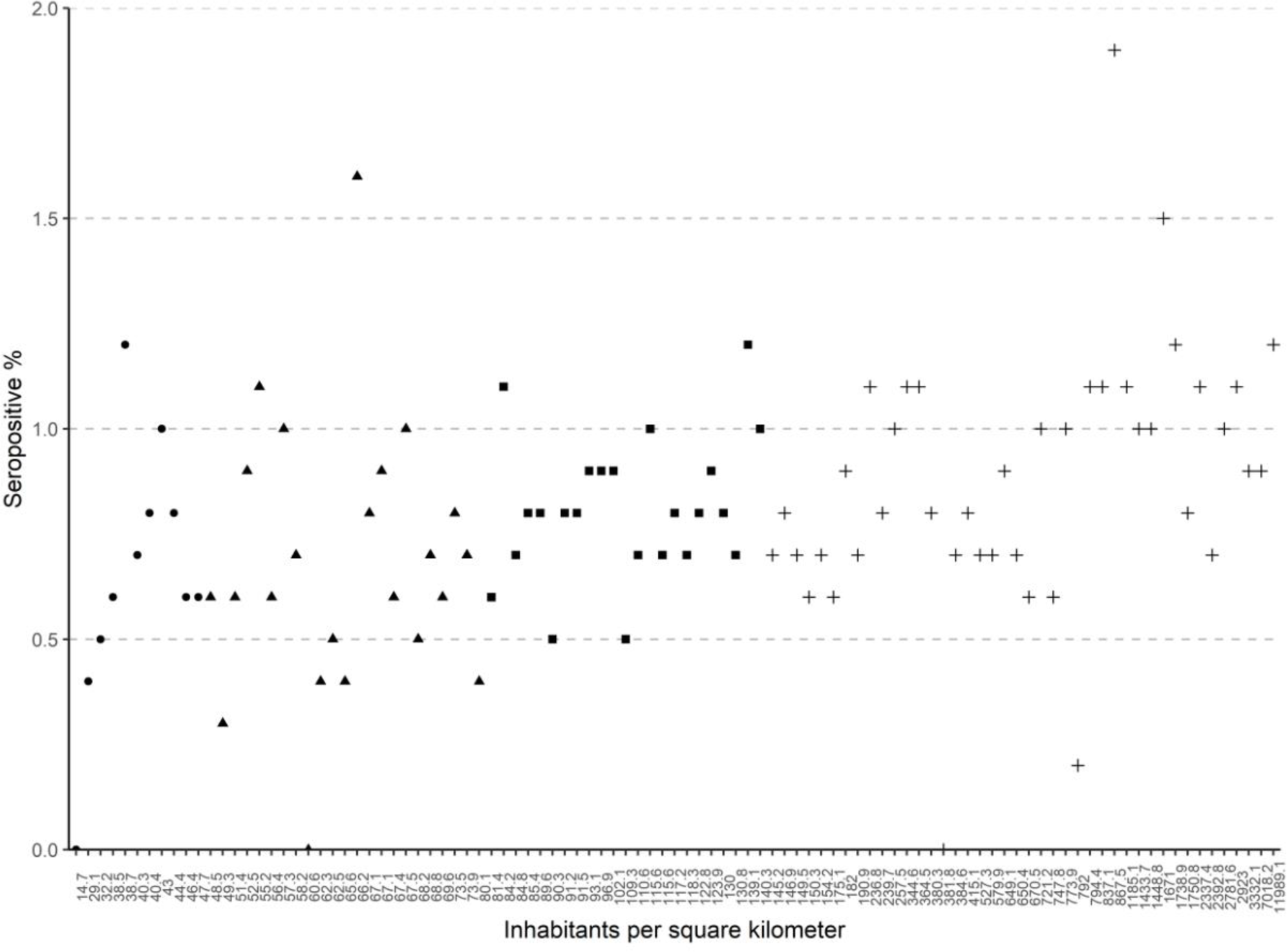
Scatterplot of seropositive % according to population density for all municipalities. Each x-axis tick represents a municipality with the corresponding population density. Dots, triangles, squares, and crosses represent population density quartiles from lowest to highest quartile, respectively.

**Supplementary figure 10:**
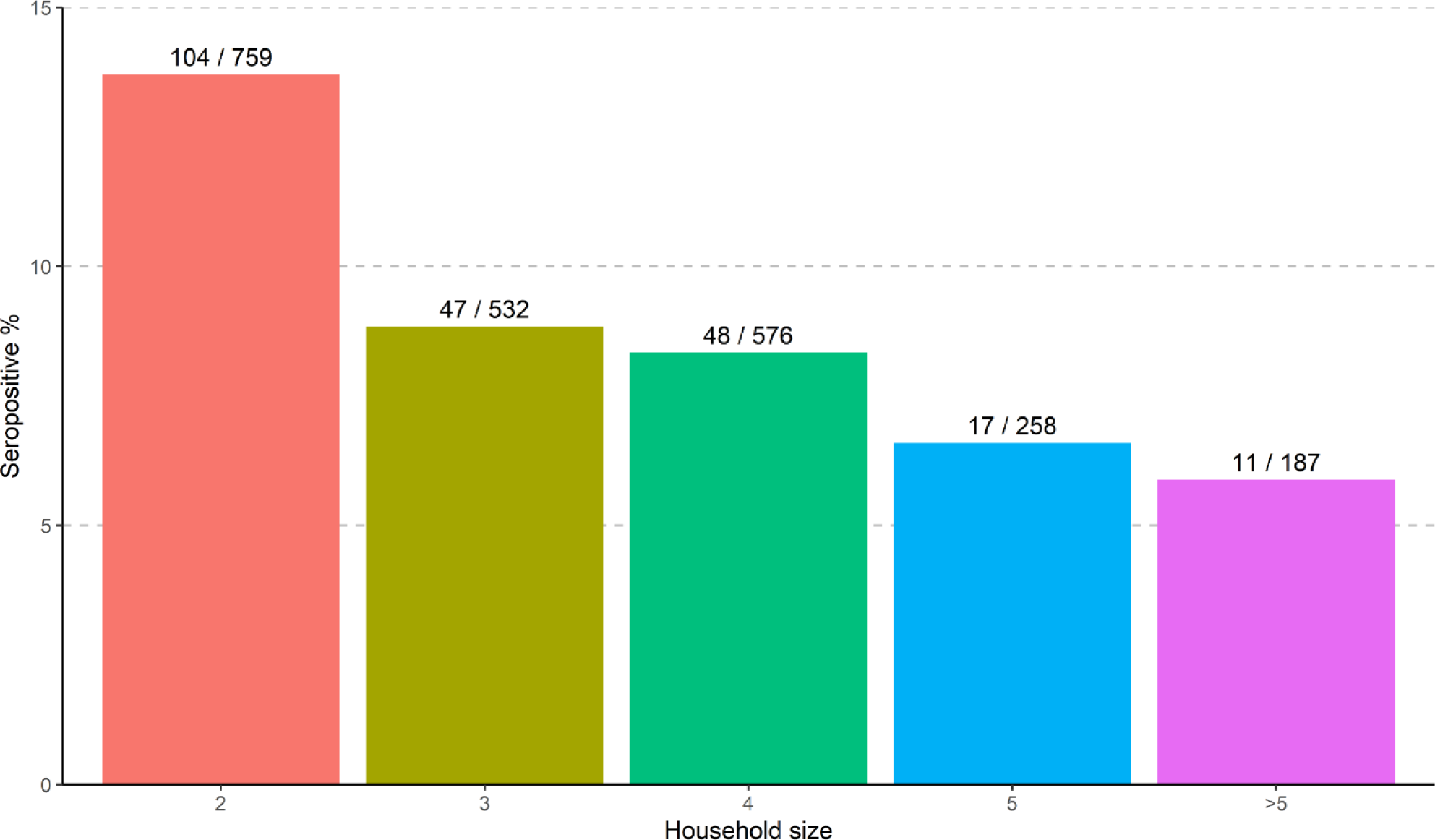
SARS-CoV-2 seropositive % in households with a COVID-19 infected person stratified by household size. Numbers above bars represent number of seropositive and seronegative participants in each household size.

**Supplementary figure 11:**
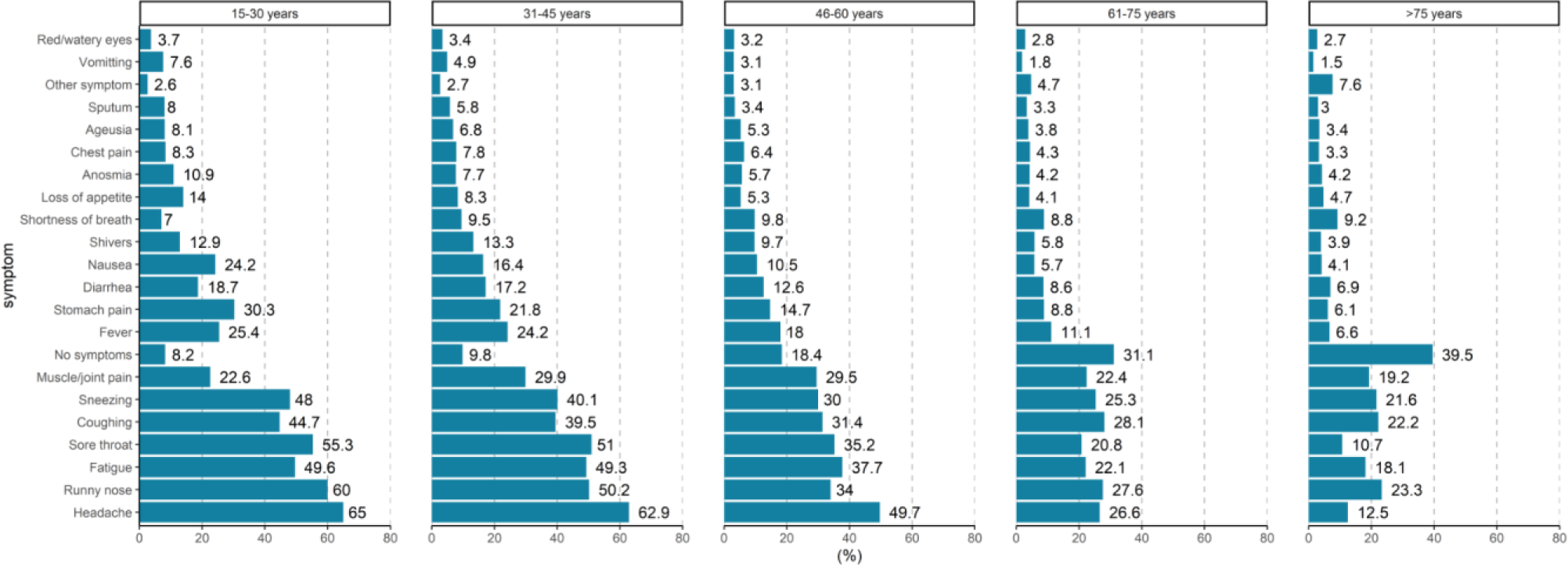
Proportion of persons who experienced symptoms stratified for age groups. Numbers next to bars represent percentages.

**Supplementary figure 12:**
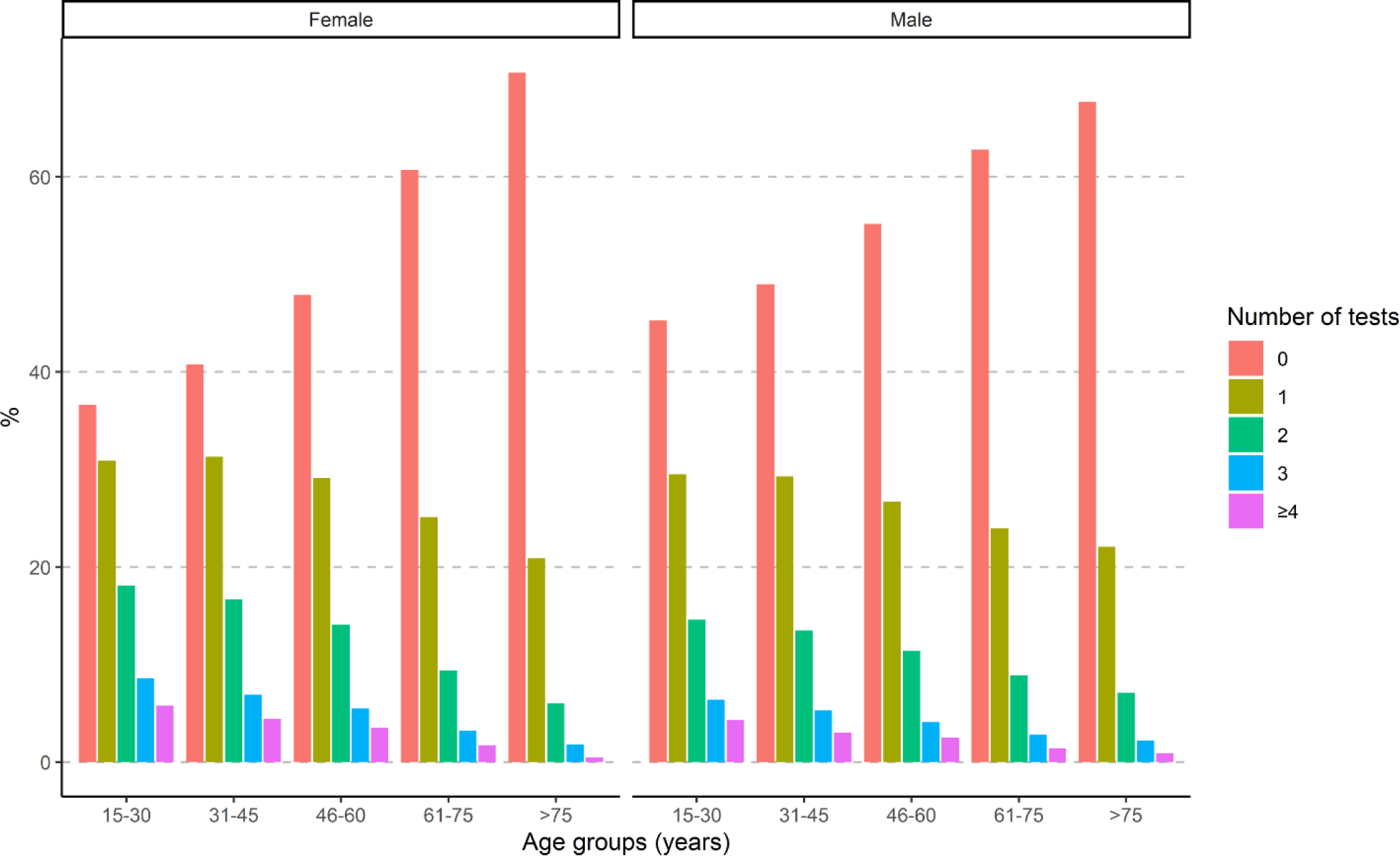
Number of previous tests for 318,552 participants stratified for age groups and sex.

## Appendix

**Table 4:** In house validation (cases=600 individuals, controls=150 individuals)

| LIVZON |  |  |  |  |  |
| --- | --- | --- | --- | --- | --- |
| Batchnummer | Expiration date for POCT | Sensitivity | 95% CI | Specificity | 95% CI |
| CK2004310410 | 14.10.2020 | 93.3% | 88.1-96.7 | 98.2% | 96.7-99.1 |
| CK2004350410 | 19.10.2020 | 92.7% | 87.3-96.3 | 97.5% | 95.9-98.6 |

## The questionnaire

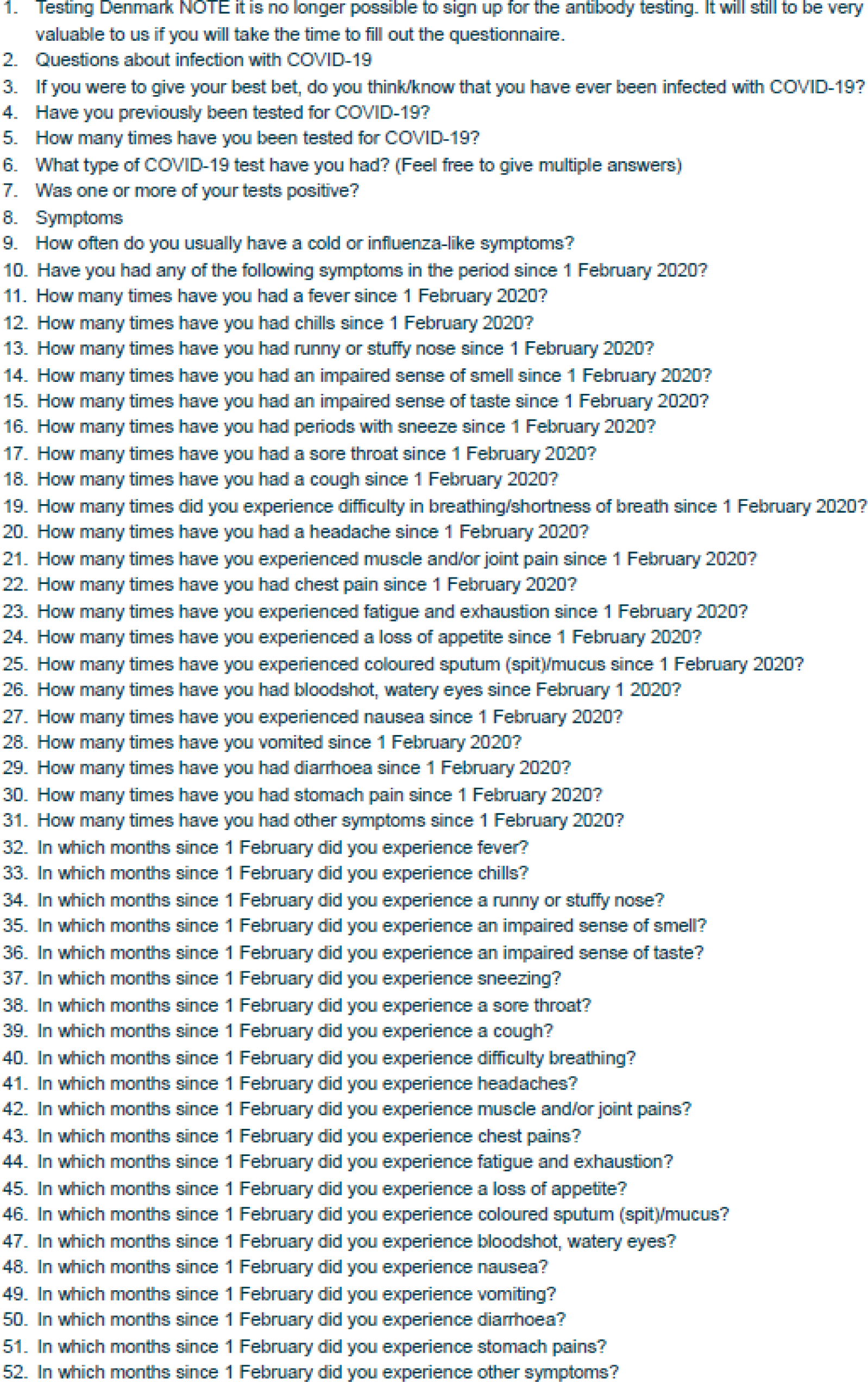
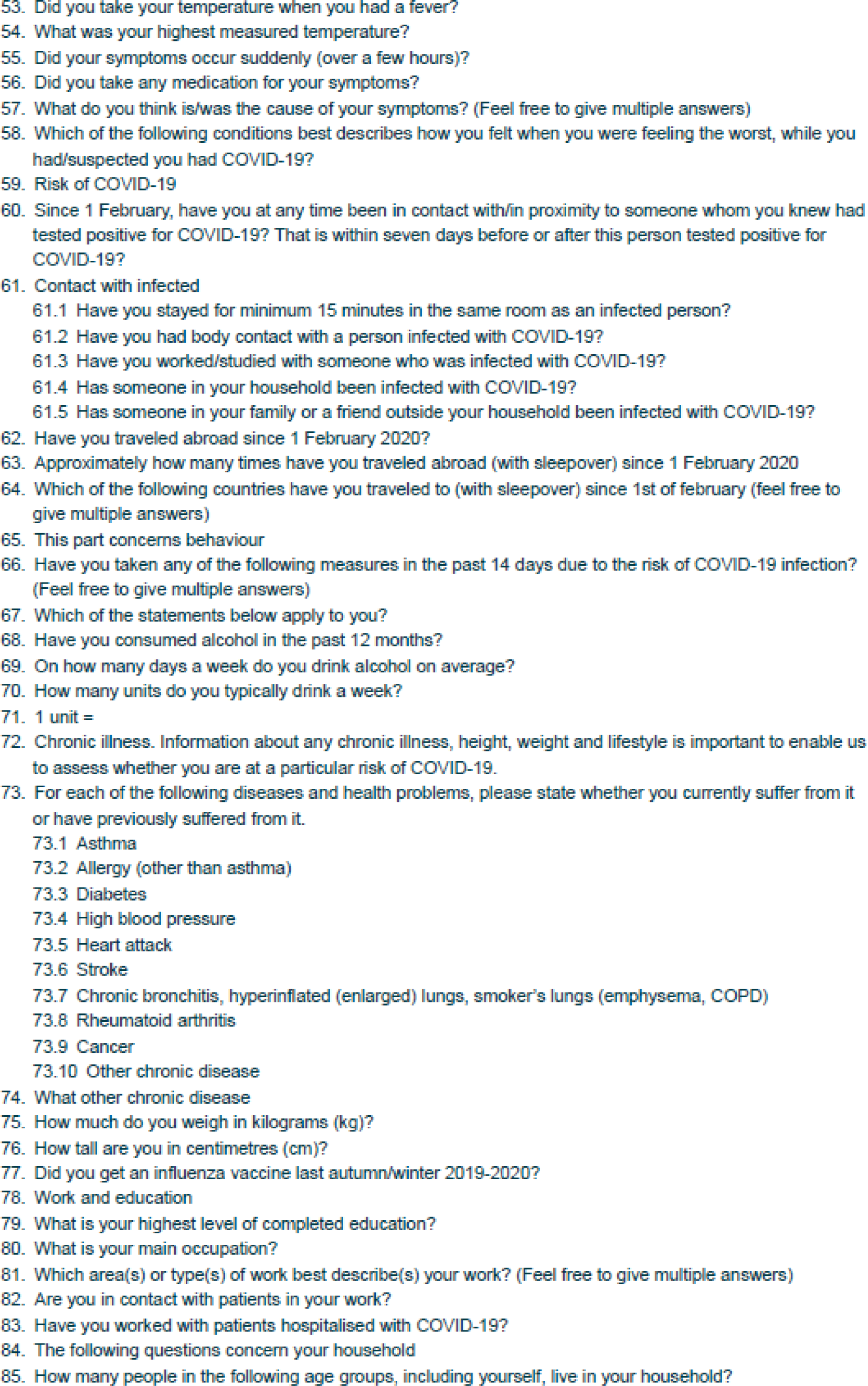
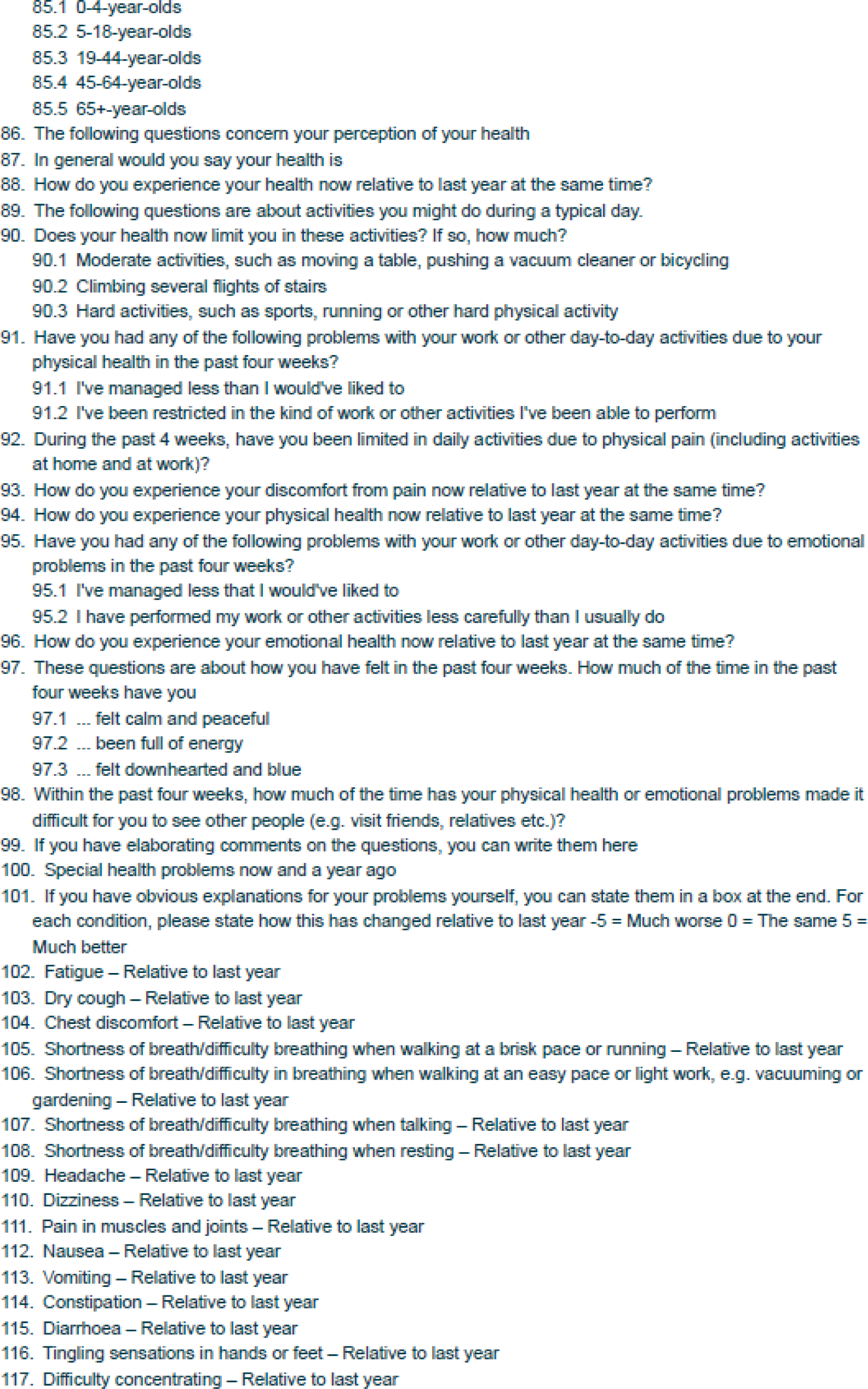
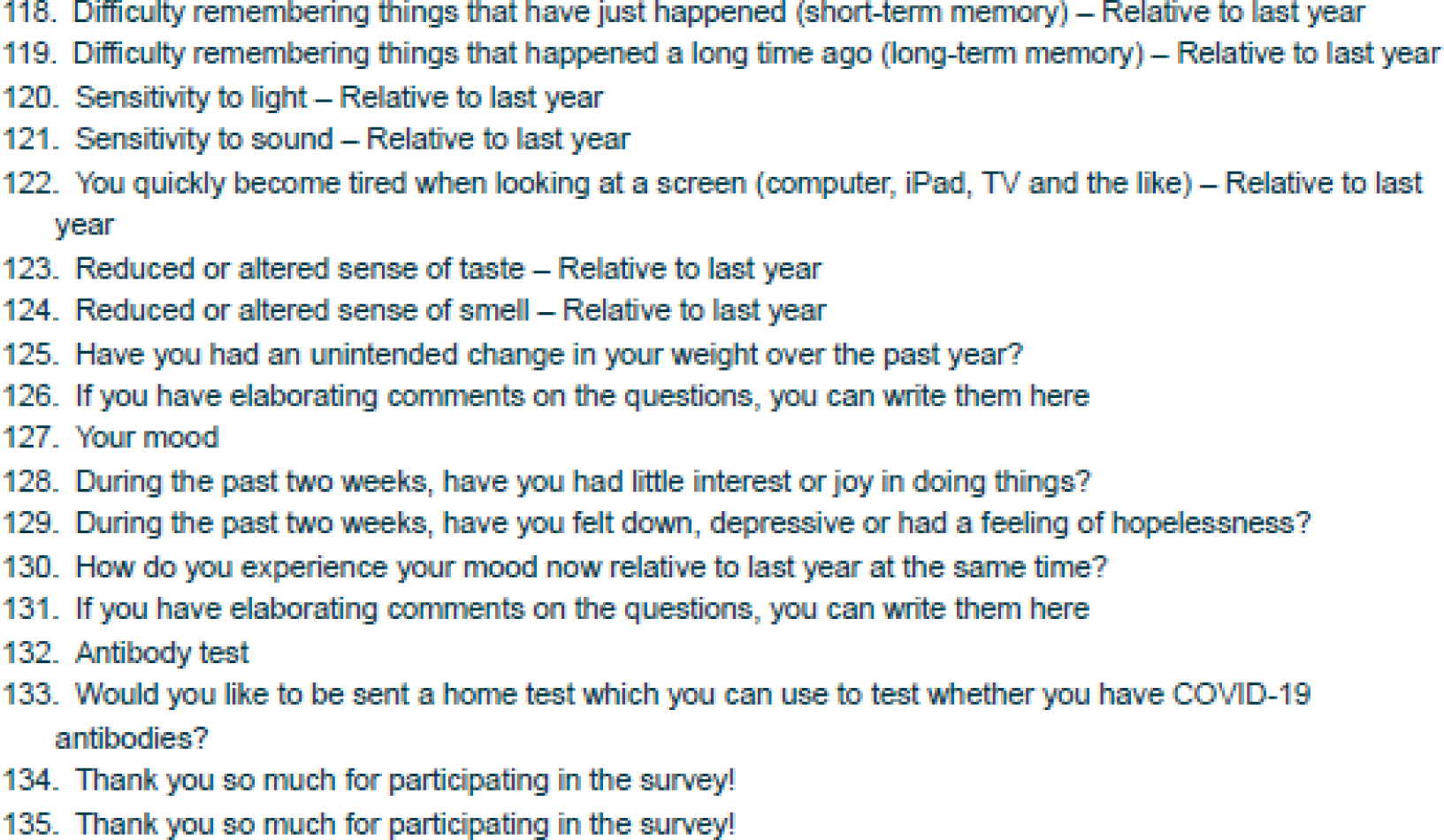

